# Emerging Therapies for COVID-19: the value of information from more clinical trials

**DOI:** 10.1101/2022.03.29.22273041

**Authors:** Stijntje W. Dijk, Eline Krijkamp, Natalia Kunst, Cary P. Gross, John B. Wong, M.G. Myriam Hunink

## Abstract

**Objectives:** The COVID-19 pandemic necessitates time-sensitive policy and implementation decisions regarding new therapies in the face of uncertainty. The aim of this study was to quantify consequences of approving therapies or pursuing further research: either immediate approval, use only in research, approval with research (e.g., Emergency Use Authorization), or reject.

**Methods:** Using a cohort state-transition model for hospitalized COVID-19 patients, we estimated quality-adjusted life years (QALYs) and costs associated with the following interventions: Hydroxychloroquine, Remdesivir, Casirivimab-Imdevimab, Dexamethasone, Baricitinib-Remdesivir, Tocilizumab, Lopinavir-Ritonavir, and Interferon beta-1a, and usual care. We used the model outcomes to conduct cost-effectiveness and value of information analyses from a US healthcare perspective and a lifetime horizon.

**Results:** Assuming a $100,000-per-QALY willingness-to-pay-threshold, only Remdesivir, Casirivimab-Imdevimab, Dexamethasone, Baricitinib-Remdesivir and Tocilizumab were (cost-) effective (incremental net health benefit 0.252, 0.164, 0.545, 0.668 and 0.524 QALYs and incremental net monetary benefit $25,249, $16,375, $54,526, $66,826 and $52,378). Our value of information analyses suggest that most value can be obtained if these 5 therapies are approved for immediate use rather than requiring additional RCTs (net value $20.6 Billion, $13.4 Billion, $7.4 Billion, $54.6 Billion and $7.1 Billion); Hydroxychloroquine (net value $198 Million) only used in further RCTs if seeking to demonstrate decremental cost-effectiveness, and otherwise rejected; and Interferon beta-1a and Lopinavir-Ritonavir are rejected (i.e., neither approved nor additional RCTs).

**Conclusions and Relevance:** Estimating the real-time value of collecting additional evidence during the pandemic can inform policymakers and clinicians about the optimal moment to implement therapies and whether to perform further research.

## Introduction

Amidst over 250 million cases worldwide, and 3,000-5,500 daily hospitalizations in the US alone, the COVID-19 pandemic represents the greatest global public health crisis since 1918.^1,2^ In the absence of known effective pharmaceutical interventions during early stages of the pandemic, many clinicians prescribed treatments off-label. Since the start of the outbreak, over 3,500 clinical trials investigating potential therapies have been registered ^3^, and new trials continue to emerge. These trials are all competing for resources and patient enrollment. Decisions on early implementation of promising treatments have been the source of substantial academic and public debate, however objective criteria for research prioritization remain absent.^4,5^

Policymakers and clinician-researchers face a difficult choice; giving Emergency Use Authorization^6^ (approval of the drug conditional on conducting more research); approval of the drug for wide-spread clinical implementation; approval of the drug only in research; or rejection of the drug based on limited existing data.^4^ A “*study, then treat or reject*” approach would optimize expected benefit by gaining more certainty about treatment effects, whereas a “*treat first, investigate later*” approach seeks to prevent lives being lost due to delayed implementation and denial of potentially beneficial treatments. However, this strategy increases the risk of harm from implementation of a possibly ineffective or deleterious treatment.

At a given point in time, findings from completed clinical trials – representing the current body of evidence - can be modelled to provide an estimate of the potential (health) benefits of further research or implementing findings of existing research.^7^ A key tool to quantify this cost-benefit trade-off is Value of Information analysis (VOI). VOI quantifies the value of treatment choices made with the expected evidence from additional research, compared to making the choice based on currently available information.^7–9^ VOI is increasingly applied as part of health economic evaluations^10–17^, both to aid the determination of the optimal sample size as well as to direct research efforts to where the greatest return can be expected from finite resources.^10^ Whereas meta-analyses investigate drug efficacy and effectiveness, and commonly conclude that further research is needed based on lack of statistical significance^7^, VOI results consider both current uncertainty relevant to the decision as well as the potential consequences of making decisions with and without further evidence.^16^ These VOI analyses can be used to quantify the benefit of further research, in terms of reducing uncertainty around treatment efficacy and avoiding unintended harm that would result from premature use of a therapy that turns out to be ineffective or deleterious. The benefits of further research are then balanced against possible forsaken benefits due to delayed implementation and research costs.^7^

In this study, we apply VOI to express the value of performing either further RCTs with delayed approval decisions (use only in RCTs), emergency use while performing RCTs (approval with research), or immediate approval of treatments for COVID-19 versus rejection without further research. Focusing on drug therapies for hospitalized COVID-19 patients for which large RCTs or meta-analyses have been published to date, we examined Hydroxychloroquine^18^, Remdesivir^19,20^, Casirivimab-Imdevimab^21^, Dexamethasone^22^, Baricitinib-Remdesivir^23^, Tocilizumab^24^, Lopinavir-Ritonavir^25^ and Interferon beta-1a^20^ compared to usual care. We considered this as a non-competing choice problem, since each of these drugs may be beneficial in the armamentarium of drug therapies for COVID-19. Our analysis aims to inform both treatment decisions and research prioritization decisions regarding therapies for hospitalized COVID-19 patients and demonstrate how a VOI approach can inform clinical and public health decision-making during a pandemic.

### Box 1.

Glossary of terms

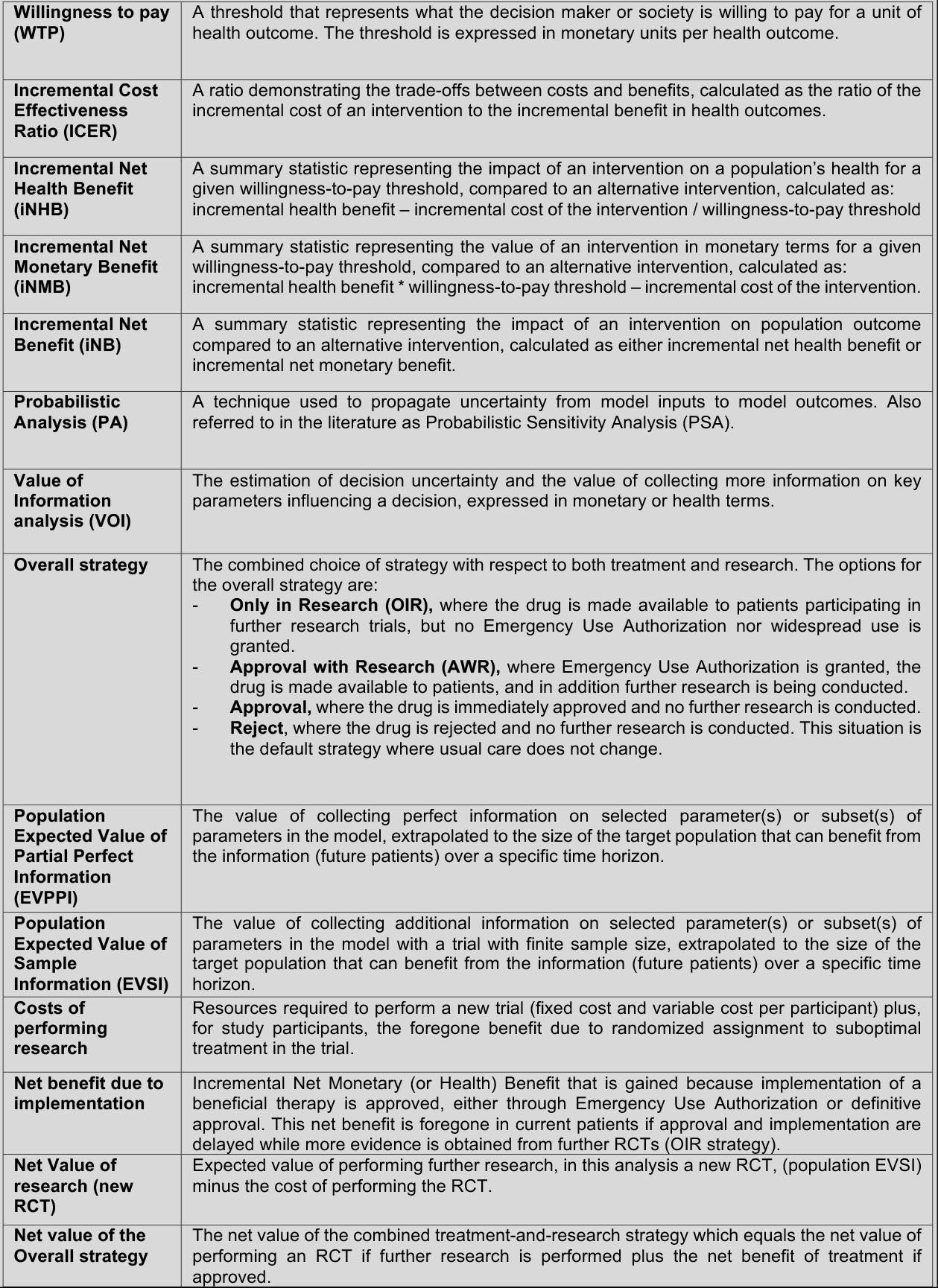

## Methods

### Decision trade-off

We performed a model-based cost-effectiveness analysis to examine both costs and health outcomes for all treatments considered. Next, we used the VOI framework to determine the net value of research. This estimate quantifies the trade-off between resources required for another RCT versus the added value of the RCT to gain more solid evidence (*Figure 1*).^26^ If the net value of research exceeds zero, performing a new RCT is worthwhile and the new evidence should be incorporated into the decision-making (*Figure 1: upper quadrants*). Ideally, trials are performed until the cost of future research exceeds the expected benefits (*lower quadrants*). This process can be applied to trials that demonstrate potential beneficial treatment effects (*right quadrants*), as well as those demonstrating no beneficial effects (*left quadrants*).

**Figure 1:**
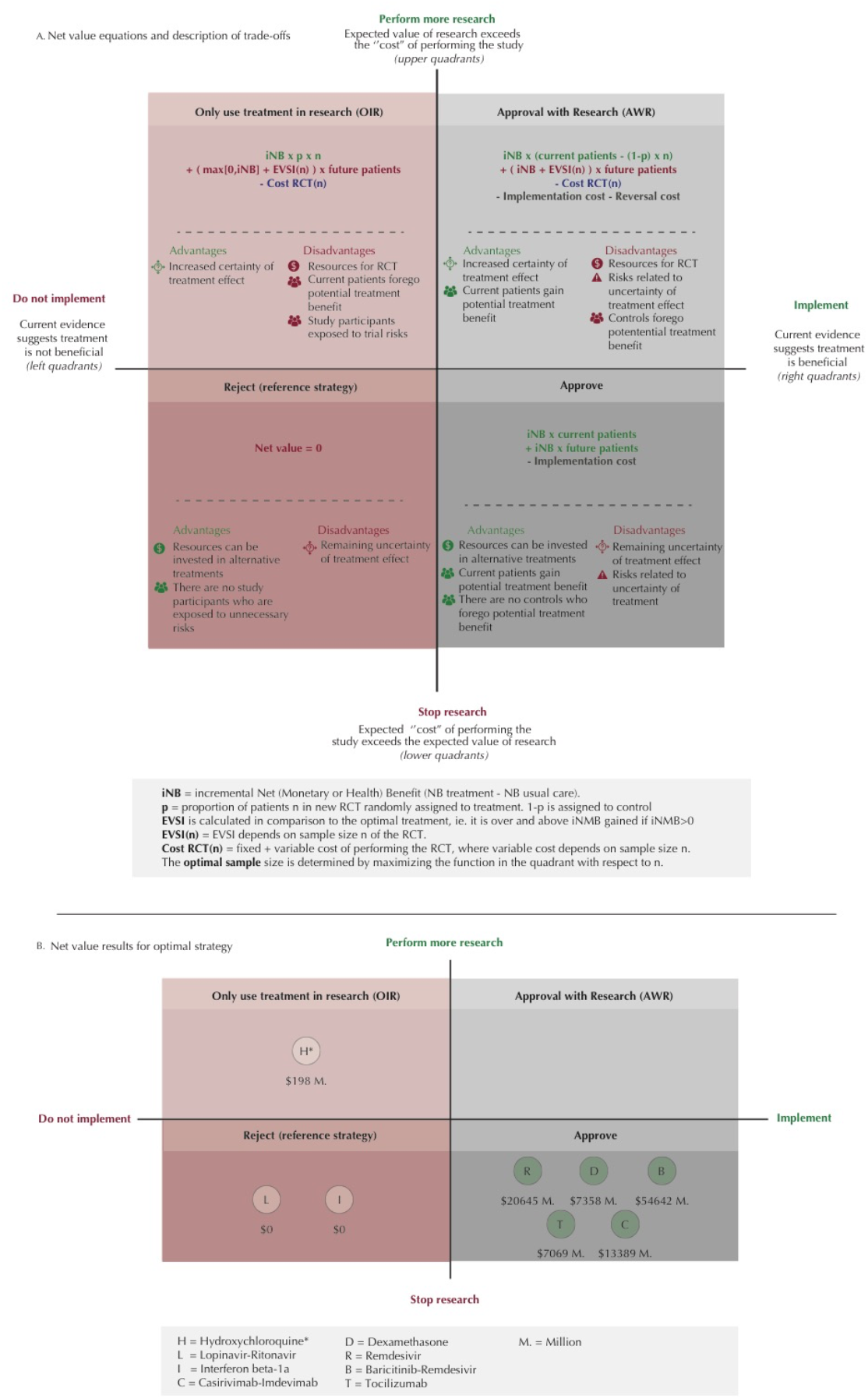
Trade-off between implementation of promising COVID-19 treatments and conducting further research. 1A: “**Net value equations and description of trade-offs”**. Demonstrates the equations used to quantify the net value for the overall strategy options, compared to reject as default strategy. These equations take iNB, EVSI, RCT cost, and number of patients (current and future) into account. iNB may be expressed in monetary units (NMB) or health units (NHB). One could also consider irrecoverable costs for the implementation of a new treatment or the possible reversal of implementation. However, in our analysis implementation and reversal costs are assumed negligible and therefor, shown in gray. The figure additionally shows the advantages and disadvantages of the corresponding implementation and research strategy. These quadrants are based on whether the drug’s current evidence suggests benefit versus standard care or placebo (the right quadrants) or not (left quadrants). Within these right and left quadrants, the upper and lower quadrants indicate whether the value of doing additional research to reduce the uncertainty in benefit exceeds the “cost” of performing additional research expressed economically or (as quality-adjusted) life years lost) in the upper quadrants or not in the lower quadrants. 1B: “**Net value results for optimal strategy**”. The net value results for the currently existing evidence and its uncertainty for eight drugs are calculated and each drug is placed in the resulting optimal health policy quadrant. Other factors, in particular ethical issues, also need to be considered to decide whether a strategy is desirable. For our study this is particularly true for Hyrdroxychloroquine. H* = Hydroxychloroquine: OIR has the highest net value if further research would demonstrate decremental cost-effectiveness (that is, saving costs but with loss of quality-adjusted life years). The ethics of investigating such decremental cost-effectiveness should be considered. If not justifiable, then hydroxychloroquine would move to the Reject category, where the net value would be 0.

### Model description

We simulated the effect of treatment for a cohort of hospitalized COVID-19 patients based on meta-analyses or large multicenter RCT’s that reported in-hospital mortality. For this simulation we used a Markov cohort state-transition model with four health states (hospitalized, recovered from hospital ward as highest level of care, recovered from intensive care unit (ICU) and dead (Appendix-Figure 1). As in the RCT’s, Hydroxychloroquine^18^, Remdesivir^19,20^, Casirivimab-Imdevimab^21^, Dexamethasone^22^, Baricitinib-Remdesivir^23^, Tocilizumab^24^, Lopinavir-Ritonavir^25^ and Interferon beta-1a^20^ were compared to the control arm (usual care) rather than to each other. We considered the implementation and research decision as a non-competing choice problem. Each drug has the potential to be a valuable component in the armamentarium against COVID-19. The various drugs have different therapeutic mechanisms, for example, as an antiviral or corticosteroid, and may be useful in sequence, in combination, or in different contexts. Similarly, from a regulatory perspective, approval is based on available safety and efficacy evidence. Since it is a non-competing choice problem, the incremental cost-effectiveness ratio (ICER) was calculated for each drug individually compared to care-as-usual. This choice allowed treatments investigated in earlier trials to become integrated in control arms in later trials. Furthermore, this modeling choice allowed for differences in context and trial populations, such as percentage of patients in the ICU and hospital wards..

All model parameters were based on best available evidence as of November 1, 2021 (Appendix-Table 1). The probabilities to transition between health states are based on a large UK cohort study^27^ of hospitalized COVID-19 patients with a mean age of 72 years. For Dexamethasone and Tocilizumab, we assumed that only patients in the ICU received treatment with these drugs. This is in accordance with current treatment recommendations.^22^ During hospitalization, patients either remained in their respective recovered states, or died from COVID-19 or other causes.^28^ The maximum hospitalization duration was 73 days.^27^ Post-hospitalization, patients were followed over their lifetime.

**Table 1:**
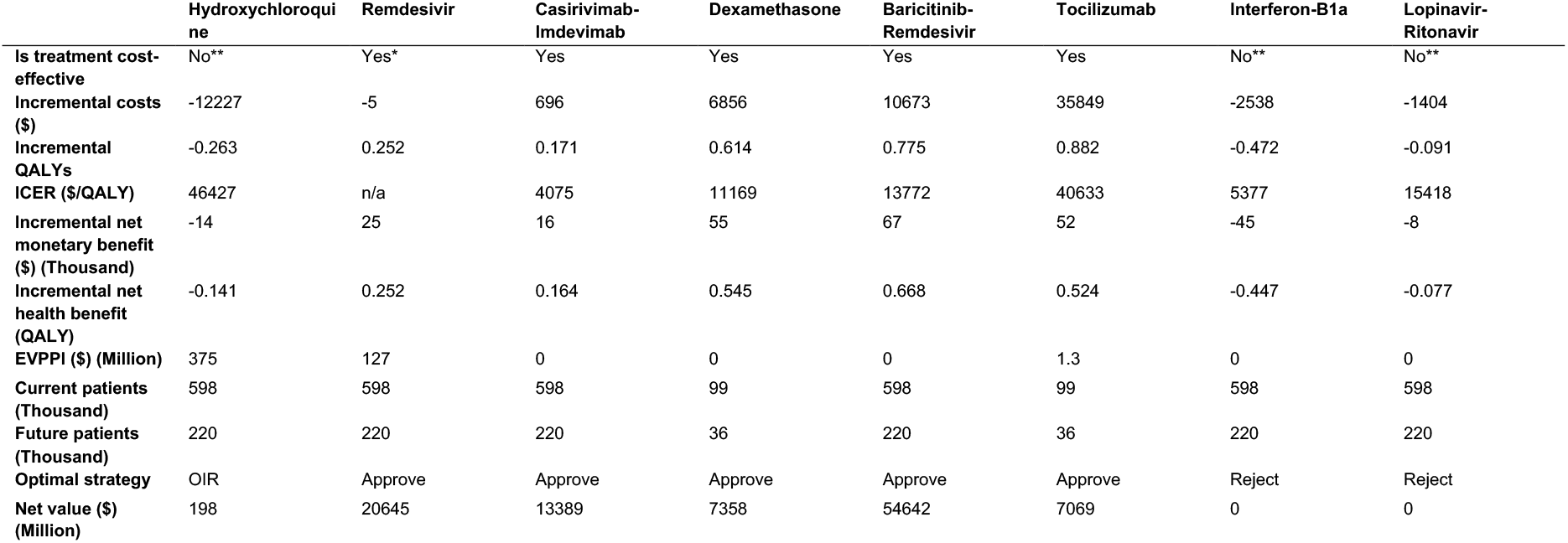
Summary results from our analysis. Results shown are the mean results from the probabilistic analysis, calculated as the treatment arm versus the care-as-usual arm of each trial at a WTP of $100,000/QALY and the results of the value of information analysis. Yes* = Treatment is dominant. Yes = Treatment is effective and ICER < WTP. No** = Treatment is cost-saving, but not enough that ICER > WTP (i.e., treatment is not decrementally cost-effective). n/a ICER not applicable because of dominance. OIR = Only in Research. Future/current patients are based on all expected hospitalized patients (Hydroxychloroquine, Remdesivir, Casirivimab-Imdevimab, Baricitinib-Remdesivir, Interferon beta-1a, Lopinavir-Ritonavir) or ICU patients only (Dexamethasone, Tocilizumab).

The model was developed in the statistical programming language R based on the DARTH framework.^29–31^ We followed the CHEERS^32^ and ISPOR^33^ reporting and analysis recommendations. We validated the model by performing internal validation using the observational cohort data^27^ and replicating the model’s cohort transitions in the decision-analytical software Amua.^34^

### Costs and effects

Health outcomes in the model were expressed as life years (LY) and quality-adjusted life-years (QALYs). Costs were estimated in 2020 US Dollars ($).^35^ The analyses were performed from a US healthcare perspective. Costs during hospitalization included daily costs per person in a hospital ward or in the ICU and depended on the estimated mean length of hospital stay based on trial data. Treatment costs were based on the price proposed by the manufacturer or public pharmacy databases. Following hospitalization, patients in the ICU Recovery state accrued a one-time rehabilitation cost based on treatment needs as estimated by the Dutch National Health Authority guidelines^36^ and converted to US prices for equivalent services. Recovered patients incurred mean healthcare costs for US citizens according to their age group annually until their death.^37^ A review of 59 novel therapeutic drugs informed the costs for additional research, where the fixed cost estimate of trials up to 26 weeks were adjusted pro rata to represent a shorter trial duration of 3 months.^38,39^ We applied a 3% annual discount rate for both costs and effects^40^, and a $100,000 willingness-to-pay (WTP) threshold.^41–43^

### Uncertainty analyses and value of information

To assess the benefit of conducting further research, VOI analysis quantifies the opportunity cost of suboptimal decisions due to uncertainty. It takes a random sample of the value of each model parameter (Appendix-Table 1 and Supplementary Excel file) and evaluates the resulting outcomes to determine the optimal strategy (treatment or usual care) for that iteration (Probabilistic Analysis, PA). We performed a PA and calculated the expected value of each strategy for each of the 10,000 iterations. Input parameter distributions were lognormal (treatment effects), beta (utilities, transition probabilities), uniform or triangular (costs, or when distributions were not available from data sources).^26^ Detailed information on all parameters is included in the Excel supplementary file.

With perfect knowledge of the parameter values chosen from their distributions for each of the 10,000 iterations of the PA, we determined the optimal strategy and its expected value for each iteration. Averaging these yielded the average expected value of the 10,000 decisions made with perfect information. Next, we calculated the difference between the average expected value of the 10,000 decisions made with perfect information and the decision based on the average expected value of each of the strategies, i.e. the decision made with current information. This difference yielded the loss in expected value due to suboptimal decisions as a result of parameter uncertainty, also known as the expected value of perfect information. We expressed the VOI results on a single scale of net monetary benefit (NMB) by converting QALYs to a monetary amount by multiplying these QALYs by a societal willingness-to-pay, e.g., $100,000 per QALY gained, and subtracting the resource costs.

Similarly, we calculated the expected added value of performing an RCT to reduce *only* the uncertainty surrounding treatment-related decrease in mortality as partial perfect information (EVPPI). For drugs with identified potential positive value of further research (EVPPI > 0), we determined the value of collecting additional information on treatment efficacy with a trial of finite sample size (expected value of sample information, EVSI). We performed an EVPPI estimation with a linear-regression meta-model and EVSI with a Gaussian approximation approach as proposed by Jalal and Alarid-Escudero.^44–46^ In this approach the opportunity loss from a suboptimal decision is approximated by a linear relation of the parameters of interest.^43^ This process was followed by a Gaussian approximation which simplifies the traditional Bayesian approach by computing the posterior mean for each of the parameters of interest (i.e., treatment efficacy in our analysis).^44^ The approximation allows for multiple correlated parameters and parameters by different sample sizes, and a wide range of univariate and multivariate non-Gaussian distributions, and is computationally substantially more efficient than traditional Bayesian updating in EVSI.

Next, we calculated the cost of further trials based on fixed and variable cost across sample sizes. As it is impossible to obtain perfect information, VOI places an upper bound on the cost of additional research aimed at reducing uncertainty.^16^ The optimal sample size of a new RCT was calculated as the size at which the net value of the optimal overall strategy is highest. Given reported concerns of insufficient trial enrollment^5^, we evaluated both the optimal and a maximum feasible sample size of 2500 patients (reported in Appendix section 1.4). Net benefit obtained with Emergency Use Authorization of treatments while performing further RCTs was determined for the expected number of patients to be hospitalized in the USA while awaiting trial results and their implementation (current patients) over 3 months. The expected value of information was extrapolated to the patient population that could benefit from new trial results, i.e., the number of patients expected to be hospitalized after the trial results are available (future patients). The number of patients was calculated as the sum of the number of daily hospitalizations forecasted by the Institute of Health Metrics and Evaluation as of November 1^st^ 2021 until March1^st^ 2022.^47^

The net value for each strategy was calculated according to the equations in **Figure 1A** Potential strategies included were Reject (*left lower quadrant*), Approve (*right lower quadrant*), Approve with Research (AWR, *right upper quadrant*), or use the drug Only in Research settings (OIR, *left upper quadrant*).^4^ Rejection without further research (*left lower quadrant*) was considered the reference (‘‘default’’) strategy (net value = 0). We assumed both the AWR and Approve strategies irrecoverable implementation and reversal costs^48^ to be $0 since treatment protocols for COVID-19 are continuously and expeditiously updated, which does not require major investments, and no fixed capital investments are required for these treatments.

In sensitivity analyses we investigated main drivers of the results by testing extreme values and consequences of underlying modeling assumptions. In addition to the analyses in the manuscript assuming a WTP-threshold of $100,000, the Appendix provides the cost-effectiveness acceptability curves for each drug and in addition illustrates how the EVPPI results depend on the WTP threshold.

### Ethics approval

Medical ethical review board approval was not required since we performed mathematical modeling and simulation using published data. No data from human participants was collected in this study

## Results

### Cost-effectiveness analysis

Table 1 summarizes key findings for each treatment strategy. Incremental costs and effectiveness (in QALYs) per individual are shown in Figure 2.

**Figure 2:**
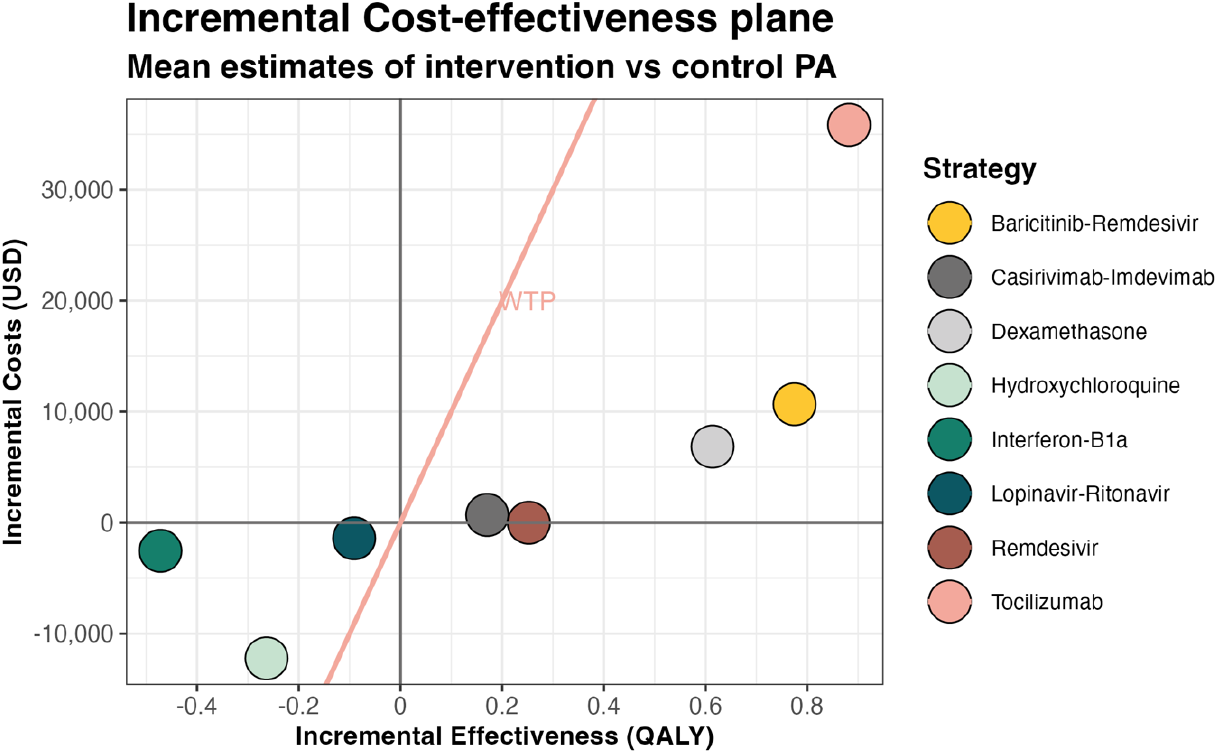
Incremental cost-effectiveness plane for mean estimates per individual resulting from the Probabilistic Analysis (PA). Incremental costs in US dollars (USD) and effects are calculated and shown as the treatment group versus the control group within the respective trial, and not in comparison to the other treatments projected. The right side of the WTP threshold line represents cost-effectiveness.

Our PA results indicate a decreased mortality and increased quality of life for Baricitinib-Remdesivir, Dexamethasone, Remdesivir and Tocilizumab, whereas Hydroxychloroquine, Interferon beta-1a and Lopinavir-Ritonavir are associated with increased mortality and decreased quality of life. Over a lifetime, the average QALY gains per patient were 0.775 [Uncertainty Interval -0.192;1.670] with Baricitinib-Remdesivir, 0.614 [0.039;1.258] with Dexamethasone, 0.252 [-0.141;0.670] with Remdesivir, 0.171 [-0.082;0.440] with Casirivimab-Imdevimab, and 0.882 [-0.052;1.937] for Tocilizumab; at an incremental cost of $10,673 [-$3930;$24372] for Baricitinib-Remdesivir, $6856 [-$19,696;$33,723] for Dexamethasone, $695 [-$33,082;$36,541] for Casirivimab-Imdevimab, and $35,849 [$20,447;$52,175] for Tocilizumab, and with marginal cost savings of -$5 [-$33,318;$235,724] for Remdesivir, making Remdesivir dominant, with higher effects and lower costs compared to usual care. Higher costs for Tocilizumab were mainly driven by treatment costs, whereas Baricitinib-Remdesivir, Dexamethasone and Casirivimab-Imdevimab costs were driven by the high healthcare costs during remaining life years in surviving patients. Conversely, lower costs for Hydroxychloroquine (-$12,227 [-$32,725;$7344]), Interferon beta-1a (-$2,538 [-$14,453;$8622]), and Lopinavir-Ritonavir (-$1,404 [-$40,584;$37,062]) were due to decreased survival leading to reduced future healthcare costs.

At a WTP of $100,000/QALY, positive incremental NMB’s were found for Baricitinib-Remdesivir ($66,826 [-$15.895;$144,126]), Dexamethasone ($54,526 [-$10,111;$120,427]), Remdesivir ($25,249 [-$23,881;$73,206]), Casirivimab-Imdevimab ($16,375 [-$25,404;$57,641]), and Tocilizumab ($52,378 [-$30,049;$149,555]), consistent with being cost-effective. The remaining strategies Hydroxychloroquine, Interferon beta-1a and Lopinavir-Ritonavir were not cost-effective. The expected values as presented above should be used to identify the optimal strategy.^49^ The uncertainty intervals reflect the range of decision uncertainty and the consequence of this uncertainty should be assessed in VOI analysis.

These PA results, uncertainty intervals, cost-effectiveness planes and cost-effectiveness acceptability curves for each treatment strategy are provided in the Appendix-Supplementary graphs and tables (Appendix-Supplementary graphs and tables –1.2/1.3/2.1-2.3/3.1-3.3/4.1/4.3/5.1-5.3/6.1-6.3/ 7.1-7.3/8.1-8.3/9.1-9.3).

### Value of information and optimal overall strategy

The population EVPPI was only positive for Hydroxychloroquine ($375 Million), Remdesivir ($127 Million) and Tocilizumab ($1.4 Million) (Table 1, Appendix Supplementary graphs and tables–1.4/1.5/2.4/3.4/4.4/5.4/6.4/7.4/8.4/9.4), suggesting further RCTs to determine treatment effect more precisely may be worthwhile. Conversely, high certainty surrounding the treatment effect of Dexamethasone and Baricitinib-Remdesivir, and the absence of benefit of Lopinavir-Ritonavir and Interferon beta-1a suggest that performing more RCTs will not affect the decision about the use of these treatments in clinical practice (Table 1). This indicates that implementing Dexamethasone ($7.4 Billion), Casirivimab-Imdevimab ($13.4 Billion), and Baricitinib-Remdesivir ($54.6 Billion), and rejecting Lopinavir-Ritonavir ($0) and Interferon beta-1a ($0) without further trials were implementation strategies with the highest overall value.

In order to decide whether further research is warranted for Hydroxychloroquine, Remdesivir, and Tocilizumab, the net value of the relevant strategies (*Figure 1B*: *upper quadrants*) was calculated for an optimal trial sample size and then compared to Reject or Approve as appropriate. The optimal overall strategy for Hydroxychloroquine, Only in Research (*Figure 1B*: *left upper quadrants, OIR*), had an estimated net value of $198 Million with an optimal sample size of 4800 patients. For the pre-set maximum feasible sample size of 2500 patients, this net value is reduced to $174 Million. The value of this further research would stem from the investigation of decremental cost-effectiveness, answering the question whether the cost-savings justify the (QA)LYs lost, which has ethical implications.

For Remdesivir and Tocilizumab, the potential benefit of further trials did not outweigh the costs of research, and the highest net value ($21 Billion and $7 Billion, respectively) was obtained with approval (*Figure 1B*: *right lower quadrant, Approve*). Our findings suggest that none of the investigated treatments should be granted Emergency Use Authorization (*right upper quadrant, AWR*).

### Sensitivity analyses

The cost-effectiveness acceptability curves illustrating the cost-effectiveness across WTP-thresholds, and tables displaying the EVPPI for different WTP thresholds are provided in the Appendix for all treatments (Appendix–Supplementary graphs and tables 2.3/2.4/3.3/3.4/4.3/4.4/5.3/5.4/6.3/6.4/7.3/7.4/8.3/8.4/9.3/9.4)

The WTP thresholds at which the therapy would be cost-effective was $0 for Remdesivir, $10,000 for Casirivimab-Imdevimab, $20,000 for Dexamethasone, $20,000 for Baricitinib-Remdesivir and $40,000 for Tocilizumab. The WTP threshold at which usual care would be cost effective was $20,000 for Lopinavir-Ritonavir, $10,000 for Interferon beta-1a, and $50,000 for Hydroxychloroquine. At lower WTP thresholds, these treatments would be decrementally cost-effective, as they would save costs through reduced long-term healthcare expenditures due to reduced survival.

## Discussion

### Summary of findings

Our results illustrate how VOI can inform policy and practice amidst a pandemic when considering whether to approve therapies, permit emergency use authorization, perform additional research or simply reject potential therapies in the treatment of hospitalized patients with COVID-19. As of November 2021, our results indicate that at a WTP of $100,000, treatment with Remdesivir, Casirivimab-Imdevimab, Dexamethasone, Baricitinib-Remdesivir and Tocilizumab leads to positive mean incremental net benefit compared to care as usual, whereas treatment with Hydroxychloroquine, Lopinavir-Ritonavir and Interferon beta-1a does not. Additionally, our results suggest sufficient certainty that decisions about treating patients with Dexamethasone, Casirivimab-Imdevimab, Baricitinib-Remdesivir, Lopinavir-Ritonavir and Interferon beta-1a would not change with further RCTs. Further research could needlessly consume resources, expose trial participants to avoidable risks and preclude them from receiving alternative (potentially) effective treatments. For Remdesivir and Tocilizumab, the net value of further trials did not outweigh the cost of research, making immediate approval their optimal overall strategies. The net value of further trials for Hydroxychloroquine outweighed the cost of research and therefore the highest net value for this drug was found in the OIR strategy. However, this further research would be conducted to investigate decremental cost-effectiveness (saving costs due to reduced survival), the ethical implications of which should be considered (see *Box 2*).

### Policy implications

As further trials unfold, the allocation of drugs in specific strategies should be considered as non-static. FDA Emergency Use Authorization has been granted for Remdesivir, Casirivimab-Imdevimab and Baricitinib-Remdesivir.^50,51^ For hospitalized COVID-19 patients, the Infectious Diseases Society of America’s (IDSA) guidelines recommend against the use of Hydroxychloroquine and Lopinavir-Ritonavir, and suggest the use of Tocilizumab, Remdesivir and Baricitinib-Remdesivir. Our findings support the IDSA’s guidelines for all investigated treatments. A summary of policy implications for each of the included treatments is provided in *Box 2*.

#### Box 2.

Policy implications and discussion per treatment

**Box 2. Policy implications and discussion per treatment**
| Treatment | Overall strategy | Current status | Reflections and other considerations |
| --- | --- | --- | --- |
| **Hydroxychloroquine** | OIR (highest net value)<br>Reject (ethical consideration) | The FDA initially granted it Emergency Use Authorization but later revoked that designation following further scientific data.<sup>52</sup> Nevertheless there are countries which continue to recommend its prescription<sup>53,54</sup> based on early studies that later received heavy criticism.<sup>55</sup> The WHO announced in June 2020 that the Hydroxychloroquine arm of the SOLIDARITY trial was stopped, based on findings from the RECOVERY and SOLIDARITY trials.<br>At least 27 trials investigating Hydroxychloroquine have been prematurely terminated and 12 withdrawn prior to enrolment<sup>3</sup>, and some countries have banned the drug in the treatment of COVID-19<sup>56</sup>. As of November 2021, still more than 100 trials were ongoing or planned to investigate (Hydroxy)chloroquine with perhaps over 100,000 participants involving all severity levels (including prophylaxis).<sup>3</sup> | Our findings support the conclusion that Hydroxychloroquine should be rejected. Yet they also suggest that there is expected positive net value in investigating Hydroxychloroquine in trials (OIR). However, the expected value from further trials stems from the drug being close to decrementally cost-effective<sup>57</sup>: it saves costs through reduced long-term healthcare expenditures due to decreased survival. On average, a healthcare cost of \$12101 per year would be “saved” by pre-mature death.<sup>37</sup> Performing further trials to demonstrate cost-savings due to reduced survival raises ethical questions to be considered.. |
| **Lopinavir-Ritonavir** | Reject | Treatment guidelines have recommended against the use of Lopinavir-Ritonavir<sup>25</sup>, after the drug showed no effect in hospitalized patients. This recommendation is in line with the findings in our analysis. According to the NMA initiative, 1 trial was reported terminated, 1 withdrawn and 1 suspended.<sup>3</sup> However, 44 trials continue to recruit patients<sup>3</sup>. | Our VOI found the highest net value in the rejection strategy. Conducting further trials may expose patients to unnecessary harms and prevents them from receiving potentially effective treatments. |
| **Interferon beta-1a** | Reject | For Interferon beta-1a some improvements of clinical aspects of COVID-19 have been identified<sup>58</sup>, however when administered in later stages of infection the drug exacerbates the disease severity due to excessive inflammation and tissue damage.<sup>58,59</sup> 3 trials were registered as terminated, 1 as withdrawn, 1 as suspended, and 54 continue recruiting.<sup>3</sup> | Our VOI found the highest net value in the rejection strategy. Conducting further trials may expose patients to unnecessary harms and prevents them from receiving potentially effective treatments. |
| **Remdesivir** | Approve | In early findings of Remdesivir in the ACTT-I trial, policymakers and regulatory agencies concluded that given the urgent need for COVID-19 treatment, the 4-day reduction in recovery time was a satisfactory proxy of the drug's effectiveness. Therefore, the FDA issued Emergency Use Authorization conditional on further research investigating its impact on mortality.<sup>60</sup> There are 29 trials investigating Remdesivir still recruiting patients as of November 2021.<sup>3</sup><br>In September 2021 a fifth large RCT, the DisCoVeRy trial<sup>61</sup> was published. The results have not yet been integrated into the major meta-analyses that we used in our analysis, but this trial also demonstrated a small but statistically non-significant beneficial effect of Remdesivir on the secondary outcome of mortality. Gyselink et al. have suggested that differences in findings across trials may be partially explained by the different levels of use of systemic steroids.<sup>62</sup> | While we found persistent uncertainty surrounding the effectiveness of Remdesivir, our model found that the benefits of widespread immediate implementation outweighed the net value of further research. This conclusion is in contrast to the WHO's recommendation to not treat with Remdesivir and to continue to recruit patients. Although both our and the WHO panels' conclusions are based on the same meta-analysis<sup>20</sup>, our findings differ because our model accounts for a lifetime horizon and the potential life years and QALYs lost, rather than statistical significance of treatment effect during the trial timeframe alone.<sup>8</sup> Further analyses that will include latest trials such as the DisCoVeRy trial<sup>61</sup>, analyze specific subgroups, and that build on network meta analyses<sup>63</sup> may help to identify the drivers of the different results for Remdesivir trials over time. |
| **Baricitinib-Remdesivir** | Approve | Baricitinib is an FDA approved treatment for rheumatoid arthritis. In November 2020 an Emergency Use Authorization was issued for the use of Baricitinib in combination with Remdesivir for hospitalized COVID-19 patients.<sup>64</sup> This guidance is based on the ACTT-2 results.<sup>23</sup> However, the ACTT-2 trial has been criticized for its inability to evaluate the effect of Baricitinib in addition to corticosteroids.<sup>64</sup> The NIH panel recommended the use of either Baricitinib or Tocilizumab in combination with Dexamethasone alone or Dexamethasone plus Remdesivir.<sup>64</sup> Their recommendations are based on the results of the COV-BARRIER study published in September 2021.<sup>64</sup> This study compares Baricitinib alone versus usual care (including corticosteroids and antivirals). As of November 2021, 11 trials are registered as recruiting patients.<sup>3</sup> | In our model, Baricitinib-Remdesivir is compared to Remdesivir as usual care, as per the ACTT-2 results.<sup>23</sup> This is made explicit, as all patients from the usual care group received Remdesivir, whereas in other trials some did and some did not. Incremental costs and acquired QALYs should therefore be interpreted as incremental to Remdesivir alone. |
| **Tocilizumab** | Approve | The treatment effect found in meta-analyses<sup>65</sup> for Tocilizumab relied heavily on the population that was treated. Tocilizumab's clinical implementation has been mainly based on the results from the REMAP-CAP<sup>24</sup> and RECOVERY<sup>65</sup> trials, and is recommended only for severe patients already receiving Dexamethasone.<sup>66</sup> As of November 2021, 28 trials in the WHO register are recruiting patients.<sup>3</sup> | In our simulation, we use the treatment effect evidence from REMAP-CAP which was solely applied to ICU patients. This is in contrast to the RECOVERY<sup>65</sup> meta-analysis, which included patients ranging from non-severe (EMPACTA<sup>67</sup>) to severe, as this reflects clinical practice. |
| **Dexamethasone** | Approve | Dexamethasone did not require Emergency Use Authorization, as it is a drug currently in use for patients who require respiratory support.<sup>68</sup> In patients with severe infections, the drug can prevent lung injury caused by community-acquired pneumonia by suppressing exuberant systematic inflammation.<sup>70</sup> In non-severe COVID-19, the use of corticosteroids is not recommended.<sup>71</sup> There are 29 trials that continue to recruit patients to investigate Dexamethasone.<sup>3</sup> | Similarly to Tocilizumab, Dexamethasone<sup>22</sup> is only given to the ICU population in our simulation, in line with the treatment recommendations.<sup>69</sup> |
| **Casirivimab-Imdevimab** | Approve | As of September 2021, the WHO issued a conditional recommendation for the use of Casirivimab and Imdevimab for only patients with seronegative status.<sup>72</sup> This recommendation is based on the statistically significant reduction of mortality in the seronegative subgroup but not the seropositive subgroup. There are 10 trials recruiting patients to investigate Casirivimab-Imdevimab. | Our current analysis included the overall effect of both seropositive and seronegative patients since our underlying population and transition probabilities represented a mixed serostatus group. Performing subgroup analysis while applying the same transition probabilities to both groups is in our opinion not justified since it would bias the results.<sup>73</sup> Future analyses investigating subgroup-specific effects with varying control group characteristics should take serostatus into account which will require the collection of subgroup-specific prognostic data. |

Although cost-effectiveness is an important tool to inform drug approval policy, there is little consensus on the WTP threshold^74^, let alone during a pandemic. A potential point of discussion is whether a WTP threshold should be higher in pandemic times due to the emergency status, or lower due to considerations of affordability when a large number of people are treated. In their assessment of the cost-effectiveness of Remdesivir, the Institute for Clinical and Economic Review applied a $50,000 WTP threshold, stating their belief that this threshold is more likely to be policy-relevant for consideration of treatments in public health emergencies.^75^ A potential additional consideration when treatment is cost-effective but raises concerns on affordability is to consider the health opportunity costs of overall budget impact of the approval of new drugs.^76,77^ The U.S. has no specified WTP threshold or a single, defined budget for healthcare spending.^74^ Historically U.S. based cost-effectiveness studies have considered ICERs at thresholds ranging from $50,000-$300,000 per QALY.^43,74,77^ Our assumption of a WTP of $100,000 follows recent criticisms in health economics research that a WTP of $50,000 would be relatively low on the basis of increases in healthcare spending and increased per capita annual income.^41–43,74^ Still, in the Appendix, we provide the cost-effectiveness acceptability curves across a wide range of WTP thresholds for all investigated treatments to inform the decision-making process. These results show that at a WTP of $50,000, the conclusions of which drugs were cost-effective (Remdesivir, Casirivimab-Imdevimab, Dexamethasone, Baricitinib-Remdesivir, and Tocilizumab) would remain the same.

### Clinical practice

For clinicians, these findings have implications for both prescription practice and participation in further trials. Firstly, our evidence-based approach can identify treatments under investigation that may not only be ineffective, but also harmful, e.g., Hydroxychloroquine is still widely prescribed based on personal beliefs or experience.^78^ With rapidly expanding evidence in the pandemic, it is paramount that clinical guidelines and practice are continuously updated to reverse previously approved therapies when appropriate so that clinicians and patients can be informed about potential benefits and harms of treatments. Furthermore, VOI summarizes existing evidence and can inform clinicians about the potential added information value, costs of further research, and ethical implications that come with it. Performing VOI can aid decision-making for policymakers, researchers, clinicians and patients on whether to initiate or continue enrollment in drug trials, stop such trials or simply approve and implement.

### Strengths and limitations

To the best of our knowledge, our study is the first to perform a VOI analysis for the treatment of COVID-19 patients. Whereas trials and meta-analyses often conclude further research is needed^7^, and where current guidelines are based on statistical significance in clinical evidence, VOI results are based on both uncertainty as well as the potential consequences of making decisions with and without further evidence. Our paper expands the potential impact of drug approval by investigating not only drug efficacy and effectiveness, but also the overall net benefits. Considering the unprecedented rollout of clinical trials investigating potential treatments for COVID-19, objective research prioritization seems paramount.^3,5^ Our model can be updated with further evidence from trials and (cumulative) meta-analyses as they become available to continuously evaluate the optimal overall strategy as the pandemic evolves.

Input parameters were based on best-available evidence. Some parameters, such as the quality of life of COVID-19 patients in 5 years and the costs of research, had to be estimated based on previous studies considering other diseases. Where necessary, we chose conservative approaches to our model parameters and settings. For example, treatment effects were only applied for reported trial duration and not extrapolated beyond and wide distributions were chosen to represent large uncertainty. Additionally, we calculated the net value for the US population alone, whereas globally more patients would benefit from determining the optimal overall strategy.

Unidentified bias in studies may, however, have affected our results. Previous papers have discussed the potential disagreements between meta-analyses and large trials.^79,80^ Single trials may not consider heterogeneity that is likely to exist across trials and centers. An advantage of single trials, however, is the more detailed group-specific information. In the case of Tocilizumab, we explicitly decided not to use the results of a meta-analysis due to the clinically relevant differences between hospital ward and ICU patients. Large network meta-analyses, such as the living WHO guideline on COVID-19 treatments,^69^ do not currently provide sufficient details to enable distinctions in treatment effect and this needs to be considered in future analyses.

With respect to the methods, our paper builds on the work by McKenna and Claxton by calculating net value and considers not only the decision for conducting further research versus implementing treatment options^7,81–83^, but also accounts for the effect of immediate implementation on patients that would have “missed out” on treatment whilst awaiting trial results.^83^ The effect of the strategy decision on current patients is especially important given the pace of hospitalizations during the pandemic. The foregone benefit of delaying approval whilst awaiting further trial results could be exceptionally high. Whilst our paper focusses on COVID-19, creating an infrastructure to investigate rapidly emerging existing trials and value of additional trials in real-time using evidence synthesis and VOI models in potential future pandemics could form the basis for informing clinical practice, research and policy decisions going forward.

The results of any VOI depend on the underlying choices and assumptions.^84^ Therefore, the limitations of our analysis need to be considered. We estimated the EVPPI with a linear-regression meta-model and EVSI with a Gaussian approximation approach as proposed by Jalal and Alarid-Escudero^44,45^. This is, however, one out of several existing estimation methods^44,45^, which differ in approaches but none of which has shown computational or statistical superiority^85^. The advantage of the chosen approximation method is the computational efficiency without introducing substantial bias^46^. A potential limitation of the linear-regression meta-model is that this normal approximation of the prior and pre-posterior distributions for parameters of interest in EVPPI and EVSI computations may introduce bias if severely non-normal.^46^ As our analysis contains parameters with sufficiently large sample sizes to approach normal distributions as per the Central Limit Theorem, we did not consider this a cause for an introduction of bias in our analysis. Finally, the current EVSI estimation methods do not consider structural uncertainty^45^, meaning that even if the true values for all input parameters are known, we are still not certain that the model reflects reality.

The key uncertainty investigated in this analysis is the estimated precision of the treatment effectiveness, as this uncertainty would be reduced by acquiring additional RCT data. Other uncertainties that could be addressed with further research include long-term morbidity and mortality, heterogeneity, adverse events of treatment, recovery time, quality of life after recovery from COVID-19, the number of hospitalizations, and costs. Furthermore, our models’ findings are not appropriate for comparing investigated treatments to each other. Our analysis does not aim to prioritize drug treatment by comparing active treatments to each other but rather focuses on the research and approval health policy questions of each studied drug regimen. Through different mechanisms and when applied in different contexts, these treatments may be useful in sequence or in combination. A head-to-head comparison of treatments based on currently available evidence would require the assumption of independent effects and comparability of study populations, which would likely strongly bias the results. Key differences in considered trials include patient populations, e.g., only ICU patients for Dexamethasone and Tocilizumab, the duration for which the treatment effect is applied, and evolving usual care as the pandemic unfolded. For example, when Remdesivir and Dexamethasone became incorporated in usual care, patients in both treatment and control arms in subsequent trials received these drugs, and accordingly our model investigated the incremental effect of the newly introduced treatment. Future network meta-analyses that identify head-to-head treatment-specific effects could provide new input for the model to help distill comparative cost-effectiveness.

Our model is based on several assumptions. The unavailability of appropriate U.S. cohort data necessitated the assumption that the large cohort of UK patients was sufficiently representative. It is likely that care for COVID-19 patients has improved since May 2020. We additionally made assumptions on the utilities of patients recovered from the ICU and Hospital ward over their lifetime. Projections of hospitalized patients^47^ may also be altered due to the rollout of vaccinations, new virus variants, (reversal of) lockdown measures, or the effect of treatment on COVID-19 transmission.

Our model did not account for age-, gender-, serostatus-, or comorbidity-specific treatment effects, and investigated reductions in mortality, but not severity of disease. Our analysis also assumed treatments were available to all patients in our simulated cohort. This assumption ignores a potential shortage of treatments in certain areas or to specific patient groups when there is a need for prioritization of resources. The analysis could, however, be repeated to investigate subgroup specific costs, effects and strategy recommendations for specific subgroups of interest.

## Conclusion

Our study demonstrates that using Hydroxychloroquine only in research; approval and implementation of Remdesivir, Casirivimab-Imdevimab, Dexamethasone, Baricitinib-Remdesivir and Tocilizumab; and the rejection of Lopinavir-Ritonavir and Interferon beta-1a provide the highest net value per November 2021. In the case of Hydroxychloroquine, this highest net value arises from decremental-cost-effectiveness, and investigating hydroxychloroquine further may be infeasible for ethical reasons. Performing ongoing VOI analyses using updated research results during the pandemic can help define the optimal moment to implement emerging therapies and whether further clinical trials are justified.

## Supporting information

Parameters

## Data Availability

Key input parameters are provided in the manuscript and Appendix. Model code, datafiles containing all input data and distributions including supplementary files will be made publicly available via [https://github.com/krijkamp/Emerging-Therapies-for-COVID-19] upon peer-reviewed journal publication.

## Author contributions

All authors had full access to data used in this study.

*Concept and design:* SD, EK, NK, MH

*Acquisition and analysis:* SD, EK, NK

*Interpretation of data:* SD, EK, NK, CG, JW, MH

*Drafting of the manuscript:* SD, CG, JW, MH

*Critical revision of the manuscript for important intellectual content:* SD, EK, NK, CG, JW, MH

*Statistical analysis:* SD, EK, NK

*Obtained funding:* SD, EK, NK, MH

*Supervision:* MH

## Data sharing statement

Key input parameters are provided in the manuscript and Appendix. Model code, datafiles containing all input data and distributions including supplementary files will be made publicly available via [https://github.com/krijkamp/Emerging-Therapies-for-COVID-19] following publication.

## Acknowledgement

The authors would like to thank the Society for Medical Decision Making COVID-19 Decision and the Gordon and Betty Moore Foundation for their support through the COVID-19 Decision Modeling Initiative. We additionally thank the Decision Analysis in R for Technologies in Health (DARTH) workgroup for providing the coding framework in R.

