## Appendix for "Emerging Therapies for COVID-19: the value of information from more clinical trials"

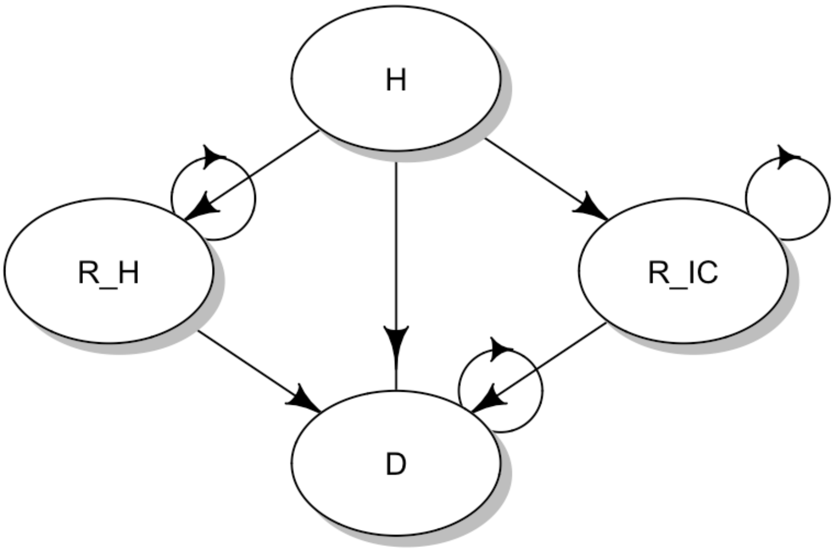

***Appendix, Figure 1***: *State-transition diagram. The state-transition diagram shows the health states applied in our model. H: Hospitalized (all patients start the simulation in the hospitalized state); R_H: Recovered from the hospital ward as the highest level of care; R_IC: Recovered from the ICU as the highest level of care; D: Dead.*

.

***Appendix, Table 1****: Key input parameters model. A full set of parameters is additionally provided in supplementary files, alongside the model code.*

| Input | | Base case | Lower bound/alpha | Upper bound/ beta | Distribution | Description | Source |
|---|---|---|---|---|---|---|---|
| **Characteristics cohort** | Age | 72 | 58 | 82 | Lognormal | Mean age of the population upon which transition probabilities are based. | (1) |
| | Probability of ICU with | | | | | | (2) |
| | Dexamethasone and Tocilizumab | 1 | 1 | 1 | NA | Dexamethasone and Tocilizumab treatment is only applied to patients who are admitted to the ICU. | |
| | All other treatments | 0.165 | 3001 | 15182 | Beta | Probability of being admitted to the ICU as the highest level of care. | (1) |
| | Mortality in | | | | | Probability of mortality if admitted to the ICU. | (1) |
| | ICU | 0.449 | 1346 | 1655 | Beta | | |
| | Hospital ward | 0.343 | 6902 | 13231 | Beta | Probability of mortality if admitted to the hospital ward. | (1) |
| **Utilities** | Quality of life in | | | | | | |
| | ICU | 0.050 | 0 | 0.250 | Triangular | Quality of life in the hospital ward, based on Health Utility Index measurements for patients hospitalized with SARS – Critical care to severe symptoms. | (3) |
| | Hospital Ward | 0.500 | 0.250 | 0.880 | Triangular | Quality of life in the hospital ward, based on Health Utility Index measurements for patients hospitalized with SARS – Severe symptoms to general population level. | (3) |
| | Recovered from ICU | 0.677 | 0.957 | 0.457 | Beta | Quality of life after recovering from the ICU, assessed prospectively in a post-ICU patient population after 5 years using the EQ-5D tool. Assumption that COVID-19 ICU patients experience the same quality of life as the general post-ICU population. | (4) |
| | Recovered from Hospital Ward | 0.880 | 41300 | 5631 | Beta | Quality of life having recovered from the hospital ward. Patients revert back to the utility of the general population, based on Medical Expenditure Panel Survey data using the EQ-5D tool. | (5) |
| **Costs** | Cost Hospital ward | 2570 | 1371 | 3552 | Triangular | Cost per day of being in the hospital ward as the highest level of care in 2020 USD. | (6) |
| | ICU with mechanical ventilation | 5209 | 4167 | 6251 | Uniform | Cost per day of being in the ICU with mechanical ventilation in 2020 USD. | (7) |
| | ICU without mechanical ventilation | 4178 | 3343 | 5015 | Uniform | Cost per day of being in the ICU without mechanical ventilation in 2020 USD. | (7) |
| | Healthcare (annual) | 12101 | 9681 | 14521 | Uniform | The mean cost of healthcare per year for the respective age group in the US | (8) |
| | Rehabilitation after ICU (one-time) | 3675 | 2940 | 4410 | Uniform | Costs estimated based on mean expected recovery needs (Dutch National Health Authority: 30 sessions physiotherapy, 5 hours occupational therapy, 4 hours dietician) and US estimated prices (Physical therapy $75/h, Occupational therapy $225/h, Dietician $70/h). | (9) |
| **Treatment effect** | Remdesivir | 0.910 | 0.790 | 1.050 | Lognormal | The **Relative Risk** of using Remdesivir versus care as usual, applied for the first 28 days of hospitalization. Based on the results of the meta-analysis reported in the SOLIDARITY trial. | (10) |
| | Dexamethasone with ventilation | 0.640 | 0.510 | 0.810 | Lognormal | The **Hazard Ratio** of using Dexamethasone versus care as usual, applied for the first 28 days of hospitalization for patients with mechanical ventilation. Based on the results reported in the RECOVERY trial. | (2) |
| | Dexamethasone without ventilation | 0.820 | 0.720 | 0.940 | Lognormal | The **Hazard Ratio** of using Dexamethasone versus care as usual, applied for the first 28 days of hospitalization for patients without mechanical ventilation. Based on the results reported in the RECOVERY trial. | (2) |
| | Hydroxychloroquine | 1.090 | 0.970 | 1.230 | Lognormal | The **Relative Risk** of using Hydroxychloroquine versus care as usual, applied for the first 28 days of hospitalization. Based on the findings of the RECOVERY trial. | (11) |
| | Interferon bèta-1a | 1.160 | 0.960 | 1.390 | Lognormal | The **Relative Risk** of using Interferon bèta-1a versus care as usual, applied for the first 28 days of hospitalization. | (10) |
| | Tocilizumab | 0.610 | 0.467 | 0.800 | Lognormal | The **Odds Ratio** of using Tocilizumab versus care as usual, applied for the first 28 days of hospitalization. Based on the REMAP-CAP trial. | (12) |

| | | | | | | | |
|---|---|---|---|---|---|---|---|
| | Lopinavir-Ritonavir | 1.030 | 0.910 | 1.170 | Lognormal | The **Relative Risk** of using Lopinavir-Ritonavir versus care as usual, applied for the first 28 days of hospitalization. Based on the findings of the RECOVERY trial. | (13) |
| | Baricitinib-Remdesivir | 0.650 | 0.390 | 1.090 | Lognormal | The **Hazard Ratio** of using Baricitinib-Remdesivir versus care as usual (Remdesivir), applied for the first 28 days of hospitalization. Based on the findings of the ACTT-2 trial. | (14) |
| | Casirivimab-Imdevimab | 0.940 | 0.860 | 1.030 | Lognormal | The **Hazard Ratio** of using Baricitinib-Remdesivir versus care as usual, applied for the first 28 days of hospitalization. Based on the findings of the RECOVERY trial. | (15) |
| **Hospitalizations** | Current patients | 597799 | | | NA | Number of patients expected to be hospitalized overall while another RCT would be conducted. Set at 3 months. Based on IHME projections, dataset downloaded 11 November 2021, projections between November 1st 2021 until March 4th 2022. In treatment options where only patients in the ICU receive treatment this number is adjusted according to the proportion of ICU hospitalizations. | (16) |
| | Future patients | 219874 | | | NA | Number of patients expected to be hospitalized excluding current patients. Based on IHME projections, dataset downloaded 11 November 2021, projections between November 1st 2021 until March 4th 2022. In treatment options where only patients in the ICU receive treatment this number is adjusted according to the proportion of ICU hospitalizations. | (16) |

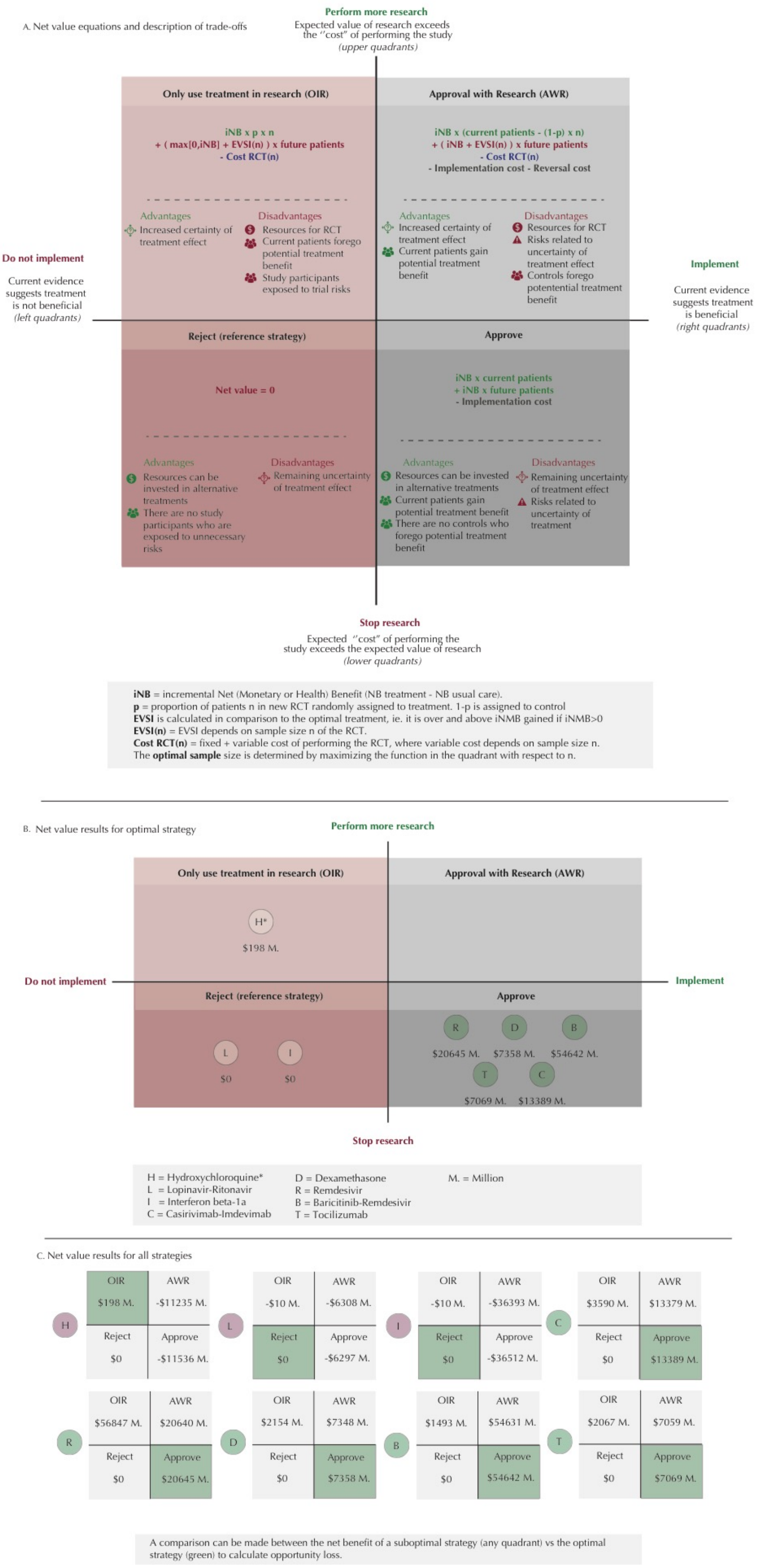

**Appendix, Figure 2:** Trade-off between implementation of promising COVID-19

treatments and conducting further research.

1A Demonstrates the equations used to quantify the net value for the overall strategy options, compared to reject as default strategy. These equations take iNB, EVSI, RCT cost, and number of patients (current and future) into account. One could also consider irrecoverable costs for the implementation of a new treatment or the possible reversal of implementation. However, in our analysis implementation and reversal costs are assumed negligible and therefor, shown in gray. The figure additionally shows the advantages and disadvantages of the corresponding implementation and research strategy. These quadrants are based on whether the drug's current evidence suggests benefit versus standard care or placebo (the right quadrants) or not (left quadrants). Within these right and left quadrants, it is considered whether the value of doing additional research to reduce the uncertainty in benefit exceeds the "cost" of performing additional research expressed economically or (quality-adjusted) life years lost (upper quadrants) or not (lower quadrants).

1B. The net value results for the currently existing evidence and its uncertainty for seven drugs are calculated and each drug is placed in the resulting optimal health policy quadrant. Other factors, in particular ethical issues, also need to be considered to decide whether a strategy is desirable. For our study this is particuallry true for Hyrdroxychloroquine. H* = Hydroxychloroquine: OIR has the highest net value if further research would demonstrate decremental cost-effectiveness (that is, saving costs but with loss of quality adjusted life years) and it should be considered whether investigating such cost-effectiveness is considered justifable. If not, then the treatment would move to the Reject category, where the net value would be 0.

1C. Comparative net values for both optimal and suboptimal overall strategies. As circumstances or policy makers may demand continued research (AWR) due to misperceived high uncertainty(17), even though immediate approval (Approve) is indicated as the optimal overall strategy, the opportunity cost of choosing a suboptimal strategy can be determined from the difference between the two strategies. For example, for Remdesivir, there is an expected loss of at least $10 Million (23024 Million – 23014 Million) when choosing the AWR strategy instead of the Approve strategy, which becomes even higher depending on the chosen suboptimal sample size

**References Appendix**

1. Docherty AB, Harrison EM, Green CA, Hardwick HE, Pius R, Norman L, et al. Features of 20 133 UK patients in hospital with covid-19 using the ISARIC WHO Clinical Characterisation Protocol: prospective observational cohort study. bmj. 2020;369.

2. RECOVERY Collaborative Group. Dexamethasone in hospitalized patients with Covid-19. N Engl J Med. 2020;

9. Zorginstituut Nederland. Zorginstituut adviseert tijdelijk ruimere vergoeding paramedische herstelzorg voor patiënten met ernstige COVID-19. zorginstituutnederland.nl. 2020 Jul 13;

10. Consortium WHOST. Repurposed antiviral drugs for COVID-19—interim WHO SOLIDARITY trial results. N Engl J Med. 2020;

### Appendix – Supplementary graphs and tables

4.4 EVPI and EVPPI for the population 22
5 Hydroxychloroquine 23
5.1 CEA results 23
5.1.1 Uncertainty interval PA results 23
5.2 Incremental CE-plane 23
5.3 CEAF 25
5.4 EVPI and EVPPI for the population 25
5.5 Expected net benefit 26
6 Interferon-B1a 27
6.1 CEA results 27
6.1.1 Uncertainty interval PA results 27
6.2 Incremental CE-plane 27
6.3 CEAF 29
6.4 EVPI and EVPPI for the population 29
7 Lopinavir-Ritonavir 30
7.1 CEA results 30
7.1.1 Uncertainty interval PA results 30
7.2 Incremental CE-plane 30
7.3 CEAF 32
7.4 EVPI and EVPPI for the population 32
8 Remdesivir 33
8.1 CEA results 33
8.1.1 Uncertainty interval PA results 33
8.2 Incremental CE-plane 33
8.3 CEAF 35
8.4 EVPI and EVPPI for the population 35
9 Tocilizumab 36
9.1 CEA results 36
9.1.1 Uncertainty interval PA results 36
9.2 Incremental CE-plane 36
9.3 CEAF 38
9.4 EVPI and EVPPI for the population 38
Probabilistic analysis – parameter distributions 39
Cohort distributions 39

### Abbreviations

This document combined the figures that are generated in the model. By having all treatment specific figures in one section

Abbreviations used: **AWR**: Approve with research; **CE**: Cost-effectiveness; **CEA**: Cost-effectiveness analysis; **CEAF**: Cost-effectiveness acceptability curve; **ENB**: Expected net benefit; **EVPI**: Expected value of perfect information; **EVPPI**: Expected value of partial perfect information; **EVSI**: Expected value of sample information; **ICER**: Incremental cost-effectiveness ration; **iLY**: Incremental life years;
**iQALY**: Incremental Quality adjusted life years; **ICU**: Intensive care unit; **LY**: Life years; **n/a**: Not applicable; **noTrt**: No treatment (care as usual); **ND**: Not dominated; **OIR**: Only in research; **PA**: Probabilistic analysis; **QALY**: Quality adjusted life years; **RCT**: Randomized control trial; **sd**: Standard deviation; **Trt**: Treatment; **WTP**: Willingness to pay.

### 1 Summary Tables

#### 1.1 Base-case

This are the treatment specific results from the base-case analysis. This means that we only used the mean value of an input parameter in the estimation. The first column indicates if the treatment is cost-effective or not. Followed by the incremental costs ($) and QALYs, where treatment is compared to no treatment. The third column gives the ICER if applicable. And the last two columns give the incremental net monetary and net heath benefit, where again treatment is compared to no treatment.

| | Is treatment cost-effective? | Incremental Costs ($) | Incremental QALYs | ICER ($/QALY) | Incremental net monetary benefit ($) | Incremental net health benefit (QALY) |
|---|---|---|---|---|---|---|
| *Casirivimab-Imdevimab* | Yes* | -7,120 | 0.169 | n/a | 24,065 | 0.241 |
| *Remdesivir* | Yes* | -8,060 | 0.254 | n/a | 33,446 | 0.334 |
| *Dexamethasone* | Yes | 6,368 | 0.604 | 10546 | 54,010 | 0.540 |
| *Baricitinib-Remdesivir* | Yes | 10,828 | 0.798 | 13568 | 68,976 | 0.690 |
| *Tocilizumab* | Yes | 35,564 | 0.869 | 40945 | 51,293 | 0.513 |
| *Interferon-B1a* | No** | -2,161 | -0.448 | 4820 | -42,680 | -0.427 |
| *Lopinavir-Ritonavir* | No** | -1,209 | -0.084 | 14379 | -7,199 | -0.072 |
| *Hydroxychloroquine* | No** | -12,363 | -0.251 | 49204 | -12,763 | -0.128 |

#### 1.2 Probabilistic analysis (PA)

This are the treatment specific results from the probabilistic analysis. This means that this are the mean results based on the several PA iterations we ran where we took the parameter uncertainty into account.

The first column indicates if the treatment is cost-effective or not. Followed by the incremental costs ($) and QALYs, where treatment is compared to no treatment. The third column gives the ICER if applicable. And the fourth and fifth columns give the incremental net monetary and net heath benefit, where again treatment is compared to no treatment.

The last three columns give the EVPPI and number of expected future and current patients. For the EVPPI estimation, only the treatment effect of the drug is evaluated. Therefore, the EVPPI value presented here gives an indication about how valuable it is to collect information about the treatment effect of this drug. More precise, the EVPPI here gives the expected values for the hypothetical scenario where we would know the exact values of the treatment effect.

| | Is treatment cost-effective? | Incremental Costs ($) | Incremental QALYs | ICER ($/QALY) | Incremental net monetary benefit ($) | Incremental net health benefit (QALY) | EVPPI | Future patients | Current patients |
|---|---|---|---|---|---|---|---|---|---|
| *Remdesivir* | Yes* | -5.499 | 0.252 | n/a | 25249 | 0.252 | 126611511 | 219874 | 597799 |
| *Casirivimab-Imdevimab* | Yes | 695.651 | 0.171 | 4075 | 16375 | 0.164 | 0 | 219874 | 597799 |
| *Dexamethasone* | Yes | 6855.757 | 0.614 | 11169 | 54526 | 0.545 | 0 | 36289 | 98663 |
| *Baricitinib-Remdesivir* | Yes | 10673.445 | 0.775 | 13772 | 66826 | 0.668 | 0 | 219874 | 597799 |
| *Tocilizumab* | Yes | 35849.422 | 0.882 | 40633 | 52378 | 0.524 | 1353077 | 36289 | 98663 |
| *Interferon-B1a* | No** | -2538.203 | -0.472 | 5377 | -44662 | -0.447 | 0 | 219874 | 597799 |
| *Lopinavir-Ritonavir* | No** | -1403.841 | -0.091 | 15418 | -7701 | -0.077 | 0 | 219874 | 597799 |
| *Hydroxychloroquine* | No** | -12226.728 | -0.263 | 46427 | -14108 | -0.141 | 375281909 | 219874 | 597799 |

#### 1.3 Cost-effectiveness plane - all treatments

In this cost-effectiveness plane the incremental differences in costs (y-axis) and health outcomes (QALYs, x-axis) between intervention and control for the different treatments are visually shown. These results are based on the PA analysis. The mean estimates of the PA analysis are shown in the figure here. The pink line shows the willingness-to-pay threshold (WTP) of 100,000$/QALY .

NOTE: The treatment specific cost-effectiveness estimates with corresponding uncertainty are given in the treatment specific sections below.

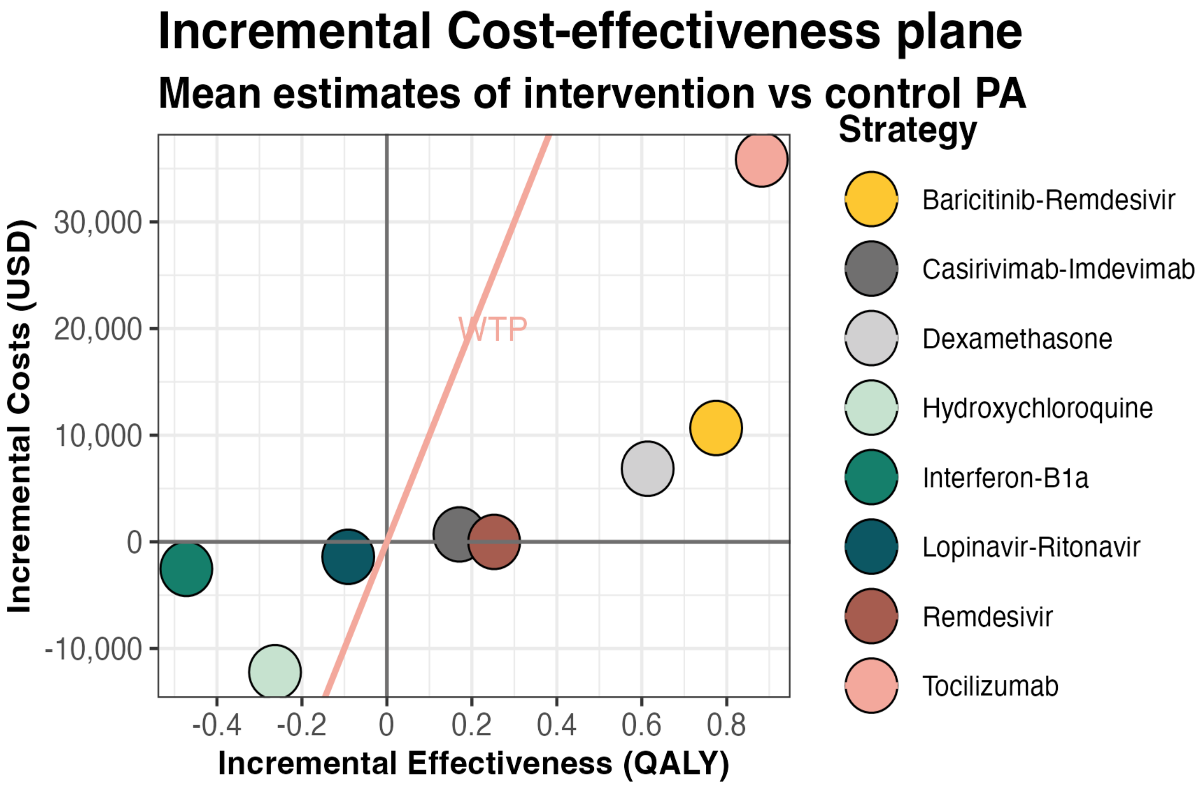

#### 1.4 Summery overview

This table gives a summary of the key outcomes (rows) of each treatment (columns). The outcomes give the results for the health outcome QALYs.

The first rows give the results from the PS analysis. Which is followed by the EVPPI values. The last rows represent the expected net value estimations for the different implementation approached. The optimal implementation strategy is given followed by the optimal sample size of a study in case the optimal strategy involves the collection of data. The net value of all strategies is given next. Having all these values can give an indication about how different the net value is for the different implementation strategies are. Since sometimes, the optimal sample size is not a feasible samples size, we also estimate the net benefit of AWR and OIR at a sample size of 2500.

| | Hydroxychloroquine | Remdesivir | Casirivimab-Imdevimab | Dexamethasone | Baricitinib-Remdesivir | Tocilizumab | Interferon-B1a | Lopinavir-Ritonavir |
|---|---|---|---|---|---|---|---|---|
| *Is treatment cost-effective?* | No** | Yes* | Yes | Yes | Yes | Yes | No** | No** |
| *Incremental Costs ($)* | -12226.728 | -5.499 | 695.651 | 6855.757 | 10673.445 | 35849.422 | -2538.203 | -1403.841 |
| *Incremental QALYs* | -0.263 | 0.252 | 0.171 | 0.614 | 0.775 | 0.882 | -0.472 | -0.091 |
| *ICER ($/QALY)* | 46427 | n/a | 4075 | 11169 | 13772 | 40633 | 5377 | 15418 |
| *Incremental net monetary benefit ($)* | -14108 | 25249 | 16375 | 54526 | 66826 | 52378 | -44662 | -7701 |
| *Incremental net health benefit (QALY)* | -0.141 | 0.252 | 0.164 | 0.545 | 0.668 | 0.524 | -0.447 | -0.077 |
| *EVPPI* | 375281909 | 126611511 | 0 | 0 | 0 | 1353077 | 0 | 0 |
| *Future patients* | 219874 | 219874 | 219874 | 36289 | 219874 | 36289 | 219874 | 219874 |
| *Current patients* | 597799 | 597799 | 597799 | 98663 | 597799 | 98663 | 597799 | 597799 |
| *Optimal strategy* | OIR | Approve | Approve | Approve | Approve | Approve | Reject | Reject |
| *Optimal sample size (N*)* | 4800 | n/a | n/a | n/a | n/a | n/a | n/a | n/a |
| *Net value for:* | n/a | n/a | n/a | n/a | n/a | n/a | n/a | n/a |
| *OIR* | 198168836 | 5684688384 | 3590068124 | 2154359630 | 14930442462 | 2066555476 | -10276460 | -10276460 |
| *AWR* | -11235187613 | 20640319686 | 13378761090 | 7348172634 | 54631440451 | 7058310458 | -36392945491 | -6307513162 |
| *Approve* | -11536116779 | 20645230711 | 13389037550 | 7358449094 | 54641716911 | 7068586918 | -36519281075 | -6297236703 |
| *Reject* | 0 | 0 | 0 | 0 | 0 | 0 | 0 | 0 |
| *Equation parameters:* | n/a | n/a | n/a | n/a | n/a | n/a | n/a | n/a |
| *Costs RCT* | 51892460 | n/a | n/a | n/a | n/a | n/a | n/a | n/a |
| *EVSI N** | 1291 | n/a | n/a | n/a | n/a | n/a | n/a | n/a |
| *Below values for sample size of N = 2500* | n/a | n/a | n/a | n/a | n/a | n/a | n/a | n/a |
| *Costs RCT N=2500* | 31951460 | n/a | n/a | n/a | n/a | n/a | n/a | n/a |
| *EVSI N=2500* | 1020 | n/a | n/a | n/a | n/a | n/a | n/a | n/a |
| *AWR N=2500* | -11326158293 | n/a | n/a | n/a | n/a | n/a | n/a | n/a |
| *OIR N=2500* | 174687339 | n/a | n/a | n/a | n/a | n/a | n/a | n/a |

NOTE: In section 1.6 the expected net value results for the health outcome LYs are given.

#### 1.5 Expected Value of Partial Perfect Information (EVPPI)

##### 1.5.1 QALY

This table gives a summary of the Expected Value of Partial Perfect Information (EVPPI) ( Million $) for the treatment parameters in each of the investigated therapies (columns) across Willingness-To-Pay thresholds (rows) health outcome quality adjusted life years (QALY).

| | Baricitinib-Remdesivir | Casirivimab-Imdevimab | Dexamethasone | Hydroxychloroquine | Interferon-B1a | Lopinavir-Ritonavir | Remdesivir | Tocilizumab |
|---|---|---|---|---|---|---|---|---|
| *0* | 0 | 0 | 0 | 0 | 0 | 0 | 245 | 0 |
| *10000* | 0 | 2 | 0 | 0 | 0 | 0 | 0 | 0 |
| *20000* | 0 | 0 | 0 | 0 | 0 | 0 | 0 | 0 |
| *30000* | 0 | 0 | 0 | 26 | 0 | 0 | 3 | 0 |
| *40000* | 0 | 0 | 0 | 283 | 0 | 0 | 12 | 66 |
| *50000* | 0 | 0 | 0 | 511 | 0 | 0 | 27 | 20 |
| *60000* | 0 | 0 | 0 | 442 | 0 | 0 | 45 | 7 |
| *70000* | 0 | 0 | 0 | 406 | 0 | 0 | 64 | 4 |
| *80000* | 0 | 0 | 0 | 387 | 0 | 0 | 84 | 2 |
| *90000* | 0 | 0 | 0 | 378 | 0 | 0 | 105 | 2 |
| *1e+05* | 0 | 0 | 0 | 375 | 0 | 0 | 127 | 1 |
| *110000* | 0 | 0 | 0 | 377 | 0 | 0 | 148 | 1 |
| *120000* | 0 | 0 | 0 | 381 | 0 | 0 | 170 | 1 |
| *130000* | 0 | 0 | 0 | 387 | 0 | 0 | 192 | 1 |
| *140000* | 0 | 0 | 0 | 395 | 0 | 0 | 214 | 1 |
| *150000* | 0 | 0 | 0 | 403 | 0 | 0 | 236 | 1 |
| *160000* | 0 | 0 | 0 | 413 | 0 | 0 | 259 | 1 |
| *170000* | 0 | 0 | 0 | 423 | 0 | 0 | 281 | 1 |
| *180000* | 0 | 0 | 0 | 434 | 0 | 0 | 303 | 1 |
| *190000* | 0 | 0 | 0 | 445 | 0 | 0 | 326 | 1 |
| *2e+05* | 0 | 0 | 0 | 457 | 0 | 0 | 348 | 1 |
| *210000* | 0 | 0 | 0 | 469 | 0 | 0 | 371 | 0 |
| *220000* | 0 | 0 | 0 | 481 | 0 | 0 | 394 | 0 |
| *230000* | 0 | 0 | 0 | 493 | 0 | 0 | 416 | 0 |
| *240000* | 0 | 0 | 0 | 506 | 0 | 0 | 439 | 0 |
| *250000* | 0 | 0 | 0 | 518 | 0 | 0 | 462 | 0 |

##### 1.5.2 Life Years

This table gives a summary of the Expected Value of Partial Perfect Information (EVPPI) ( Million $) for the treatment parameters in each of the investigated therapies (columns) across Willingness-To-Pay thresholds (rows) health outcome life years (LY).

| | Baricitinib-Remdesivir | Casirivimab-Imdevimab | Dexamethasone | Hydroxychloroquine | Interferon-B1a | Lopinavir-Ritonavir | Remdesivir | Tocilizumab |
|---|---|---|---|---|---|---|---|---|
| 0 | 0 | 0 | 0 | 0 | 0 | 0 | 245 | 0 |
| 10000 | 0 | 1 | 0 | 0 | 0 | 0 | 0 | 0 |
| 20000 | 0 | 0 | 0 | 0 | 0 | 0 | 0 | 0 |
| 30000 | 0 | 0 | 0 | 166 | 0 | 0 | 8 | 39 |
| 40000 | 0 | 0 | 0 | 511 | 0 | 0 | 25 | 7 |
| 50000 | 0 | 0 | 0 | 424 | 0 | 0 | 47 | 3 |
| 60000 | 0 | 0 | 0 | 385 | 0 | 0 | 72 | 2 |
| 70000 | 0 | 0 | 0 | 367 | 0 | 0 | 97 | 1 |
| 80000 | 0 | 0 | 0 | 362 | 0 | 0 | 124 | 1 |
| 90000 | 0 | 0 | 0 | 362 | 0 | 0 | 150 | 1 |
| 1e+05 | 0 | 0 | 0 | 367 | 0 | 0 | 177 | 1 |
| 110000 | 0 | 0 | 0 | 375 | 0 | 0 | 205 | 0 |
| 120000 | 0 | 0 | 0 | 385 | 0 | 0 | 232 | 0 |
| 130000 | 0 | 0 | 0 | 396 | 0 | 0 | 260 | 0 |
| 140000 | 0 | 0 | 0 | 408 | 0 | 0 | 287 | 0 |
| 150000 | 0 | 0 | 0 | 420 | 0 | 0 | 315 | 0 |
| 160000 | 0 | 0 | 0 | 433 | 0 | 0 | 343 | 0 |
| 170000 | 0 | 0 | 0 | 447 | 0 | 0 | 371 | 0 |
| 180000 | 0 | 0 | 0 | 461 | 0 | 0 | 399 | 0 |
| 190000 | 0 | 0 | 0 | 475 | 0 | 0 | 427 | 0 |
| 2e+05 | 0 | 0 | 0 | 490 | 0 | 0 | 456 | 0 |
| 210000 | 0 | 0 | 0 | 505 | 0 | 0 | 484 | 0 |
| 220000 | 0 | 0 | 0 | 520 | 0 | 0 | 512 | 0 |
| 230000 | 0 | 0 | 0 | 535 | 0 | 0 | 541 | 0 |
| 240000 | 0 | 0 | 0 | 551 | 0 | 0 | 569 | 0 |
| 250000 | 0 | 0 | 0 | 566 | 0 | 0 | 598 | 0 |

#### 1.6 Expected net value - LY

These values represent the expected net value estimations for the different implementation approached for the health outcome life years (LY). The optimal implementation strategy is given in the first row. The second row gives the optimal sample size of a study, in case the optimal strategy involves the collection of data. The net value of all strategies is given next. Having all these values can give an indication about how different the net value is for the different implementation strategies are. Since sometimes, the optimal sample size is not a feasible samples size, we also estimate the net benefit of AWR and OIR at a sample size of 2500.

| | Hydroxychloroquine | Remdesivir | Baricitinib-Remdesivir | Casirivimab-Imdevimab | Dexamethasone | Tocilizumab | Interferon-B1a | Lopinavir-Ritonavir |
|---|---|---|---|---|---|---|---|---|
| *Optimal strategy* | OIR | AWR | Approve | Approve | Approve | Approve | Reject | Reject |
| *Optimal sample size (N*)* | 4500 | 2500 | NA | NA | NA | NA | NA | NA |
| *Net value for:* | NA | NA | NA | NA | NA | NA | NA | NA |
| *OIR* | 167636260 | 6857202163 | 18534977308 | 4359304403 | 3426593528 | 3896483552 | -10276460 | -10276460 |
| *AWR* | -16066014778 | 24768363738 | 67738014085 | 16193360282 | 11506444880 | 13041478677 | -44558784765 | -7880767087 |
| *Approve* | -16383860973 | 24755528350 | 67748290544 | 16203636741 | 11516721339 | 13051755137 | -44735360922 | -7870490627 |
| *Reject* | 0 | 0 | 0 | 0 | 0 | 0 | 0 | 0 |
| *Equation parameters:* | NA | NA | NA | NA | NA | NA | NA | NA |
| *Costs RCT* | 49291460 | 31951460 | NA | NA | NA | NA | NA | NA |
| *EVSI N** | 1192 | 376 | NA | NA | NA | NA | NA | NA |
| *Below values for sample size of N = 2500* | NA | NA | NA | NA | NA | NA | NA | NA |
| *Costs RCT N=2500* | 31951460 | NA | NA | NA | NA | NA | NA | NA |
| *EVSI N=2500* | 928 | NA | NA | NA | NA | NA | NA | NA |
| *AWR N=2500* | -16186694003 | NA | NA | NA | NA | NA | NA | NA |
| *OIR N=2500* | 147074068 | NA | NA | NA | NA | NA | NA | NA |

NOTE: The summary overview in section 1.4 gives the expected net value results for the health outcome QALYs.

### 2 Baricitinib-Remdesivir

#### 2.1 CEA results

When reading the CEA results, carefully check if treatment or no treatment is shown as the reference. This has a major impact on the interpretation of the results.

| Outcome | Strategy | Cost | Effect | Inc_Cost | Inc_Effect | ICER | Status |
|---|---|---|---|---|---|---|---|
| Basecase | | | | | | | |
| QALY | notrt | 108318.3 | 6.062883 | NA | NA | NA | ND |
| QALY | trt | 119145.9 | 6.860919 | 10827.60 | 0.7980359 | 13567.81 | ND |
| PSA | | | | | | | |
| QALY | notrt | 109843.6 | 6.217861 | NA | NA | NA | ND |
| QALY | trt | 120517.0 | 6.992853 | 10673.45 | 0.7749926 | 13772.32 | ND |
| LY | notrt | 109843.6 | 7.318359 | NA | NA | NA | ND |
| LY | trt | 120517.0 | 8.253643 | 10673.45 | 0.9352836 | 11411.99 | ND |

##### 2.1.1 Uncertainty interval PA results

| | iLY | iQALY | iCosts | LY iNMB | QALY iNMB | LY iNHB | QALY iNHB |
|---|---|---|---|---|---|---|---|
| mean | 0.9352836 | 0.7749926 | 10673.445 | 82854.91 | 66825.81 | 0.8285491 | 0.6682581 |
| sd | 0.5580618 | 0.4666187 | 7079.587 | 49203.22 | 40141.06 | 0.4920322 | 0.4014106 |
| 2.5% | -0.2338972 | -0.1916076 | -3930.333 | -19577.14 | -15894.57 | -0.1960000 | -0.1590000 |
| 97.5% | 1.9761217 | 1.6695956 | 24371.515 | 174937.56 | 144126.15 | 1.7490000 | 1.4410000 |

#### 2.2 Incremental CE-plane

Incremental cost-effectiveness plane, shown as the incremental effectiveness (x-axis) and incremental cost ($) (y-axis). Iterations are represented by the small transparent dots, the mean is indicated by the large solid circle representing the results for QALY's and LY's separately. The dotted ellipses represent the 95% Credibility Intervals.

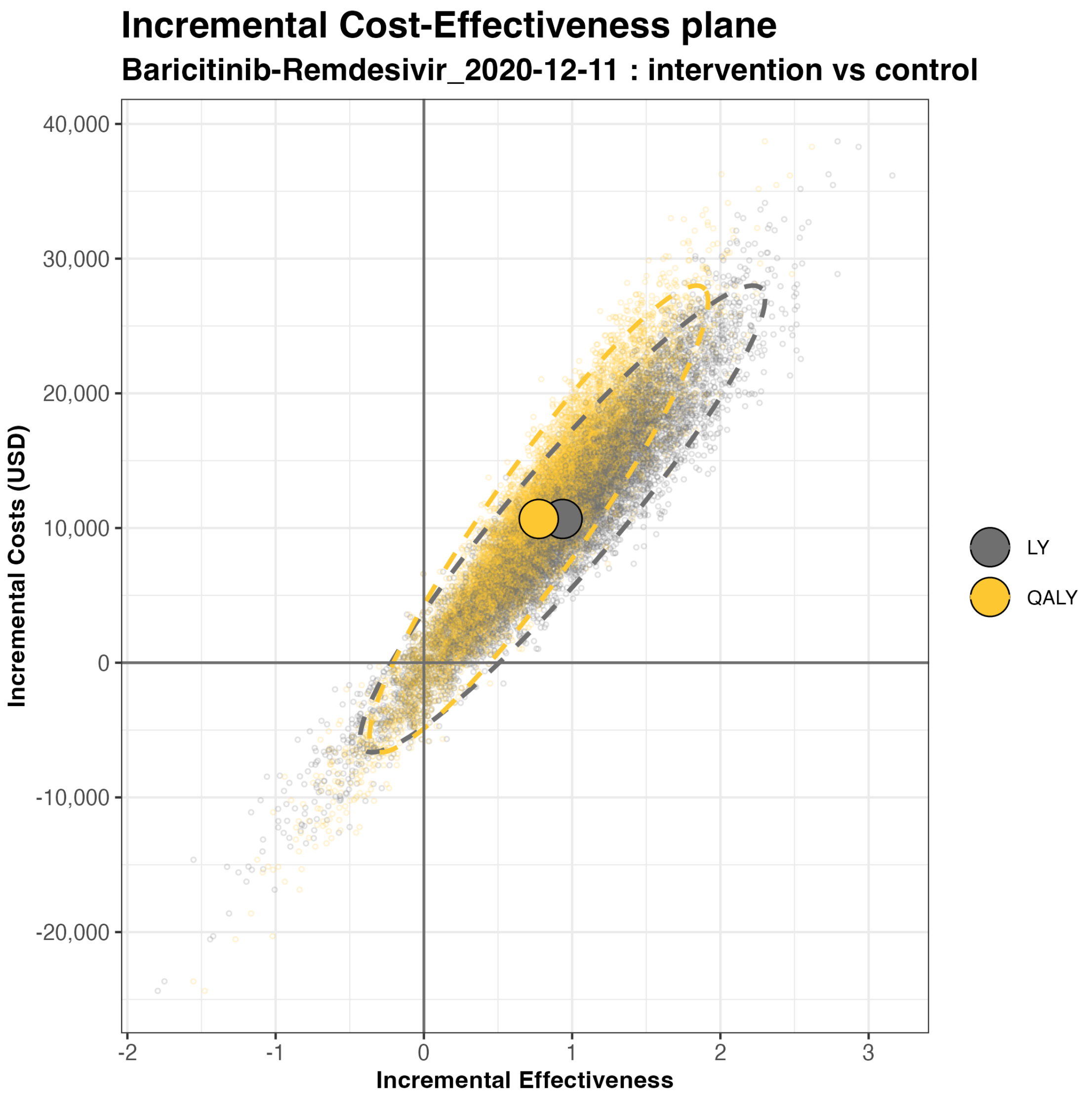

#### 2.3 CEAF

The figures show the cost-effectiveness acceptability curves (CEAF) with on the x-axes the willingness to pay thresholds expressed using (thousand $/QALY) in the top figure and (thousand $/LY) in the bottom figure. The y-axes represent the probability of the treatment and no treatment strategies being cost-effective.

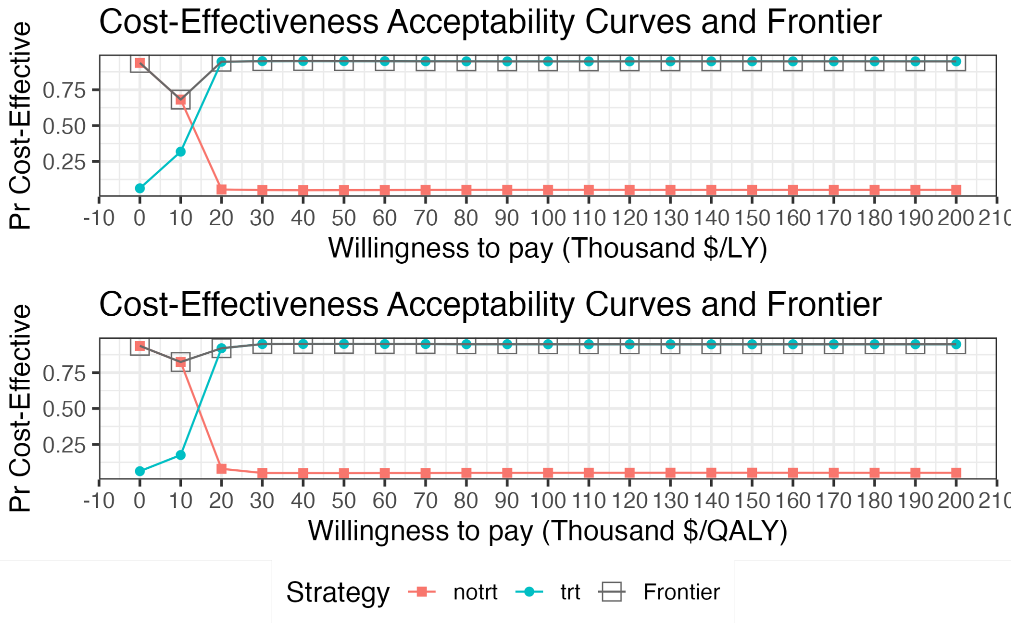

#### 2.4 EVPI and EVPPI for the population

The top panel shows the expected value of perfect (EVPI) information. The bottom panel the expected value of partial perfect (EVPPI) information.

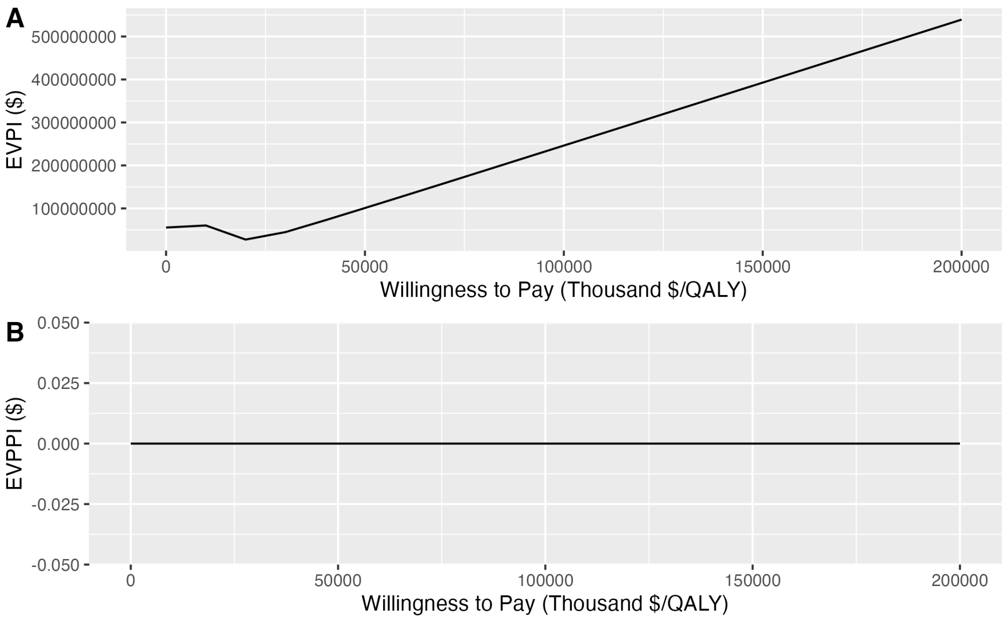

### 3 Casirivimab-Imdevimab

#### 3.1 CEA results

When reading the CEA results, carefully check if treatment or no treatment is shown as the reference. This has a major impact on the interpretation of the results.

| Outcome | Strategy | Cost | Effect | Inc_Cost | Inc_Effect | ICER | Status |
|---|---|---|---|---|---|---|---|
| Basecase | | | | | | | |
| QALY | trt | 127408.2 | 6.229414 | NA | NA | NA | ND |
| QALY | notrt | 134528.2 | 6.059959 | NA | NA | NA | D |
| PSA | | | | | | | |
| QALY | notrt | 136428.2 | 6.215980 | NA | NA | NA | ND |
| QALY | trt | 137123.9 | 6.386682 | 695.6505 | 0.170702 | 4075.234 | ND |
| LY | notrt | 136428.2 | 7.327575 | NA | NA | NA | ND |
| LY | trt | 137123.9 | 7.532699 | 695.6505 | 0.205124 | 3391.366 | ND |

##### 3.1.1 Uncertainty interval PA results

| | iLY | iQALY | iCosts | LY iNMB | QALY iNMB | LY iNHB | QALY iNHB |
|---|---|---|---|---|---|---|---|
| mean | 0.2051240 | 0.1707020 | 695.6505 | 19816.75 | 16374.55 | 0.1981675 | 0.1637455 |
| sd | 0.1590312 | 0.1324070 | 17824.8110 | 22088.49 | 21132.45 | 0.2208849 | 0.2113245 |
| 2.5% | -0.0994191 | -0.0824684 | -33082.8248 | -23428.01 | -25403.92 | -0.2340000 | -0.2540000 |
| 97.5% | 0.5248223 | 0.4401583 | 36541.1072 | 63302.18 | 57640.96 | 0.6330000 | 0.5760000 |

#### 3.2 Incremental CE-plane

Incremental cost-effectiveness plane, shown as the incremental effectiveness (x-axis) and incremental cost ($) (y-axis). Iterations are represented by the small transparent dots, the mean is indicated by the large solid circle representing the results for QALY's and LY's separately. The dotted ellipses represent the 95% Credibility Intervals.

#### Incremental Cost-Effectiveness plane

##### Casirivimab-Imdevimab_2021_06_16 : intervention vs control

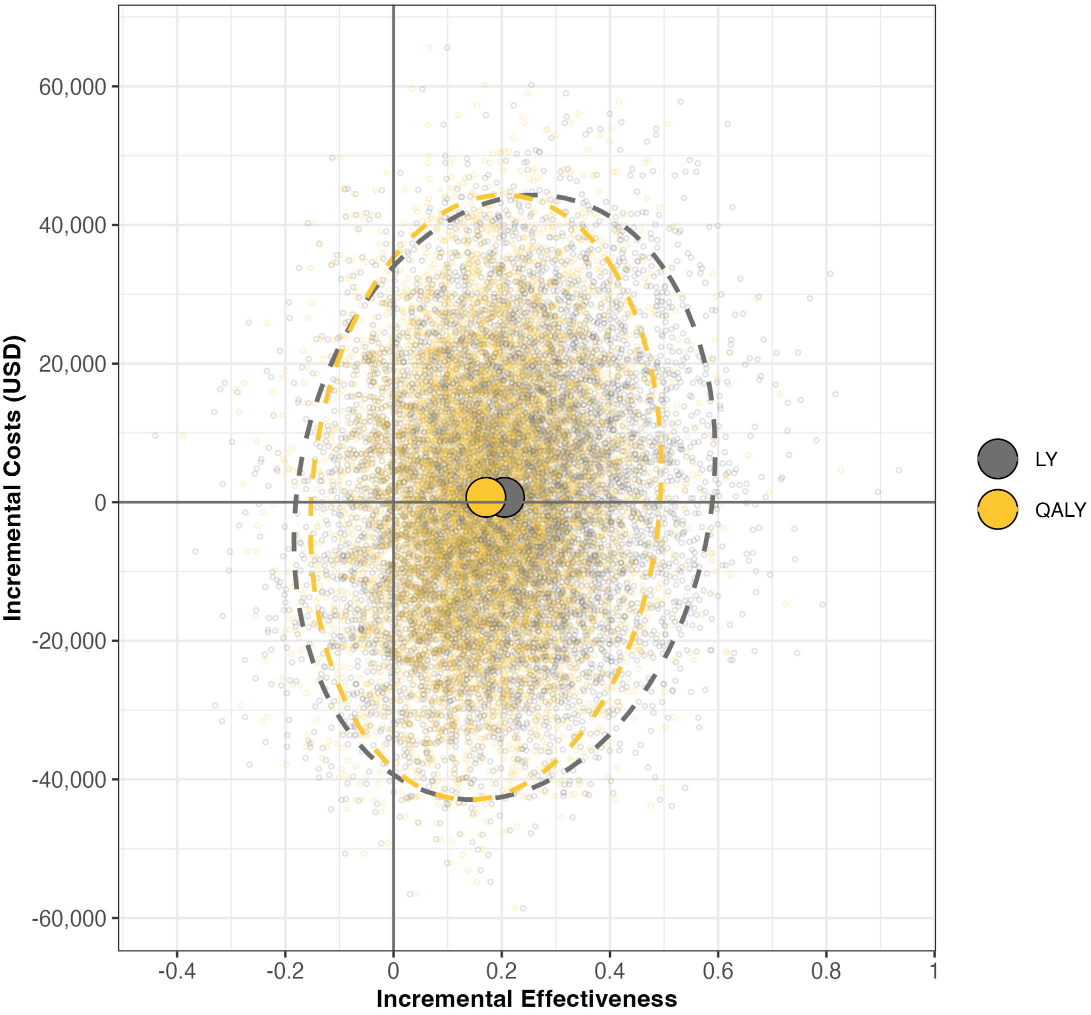

#### 3.3 CEAF

The figures show the cost-effectiveness acceptability curves (CEAF) with on the x-axes the willingness to pay thresholds expressed using (thousand $/QALY) in the top figure and (thousand $/LY) in the bottom figure. The y-axes represent the probability of the treatment and no treatment strategies being cost-effective.

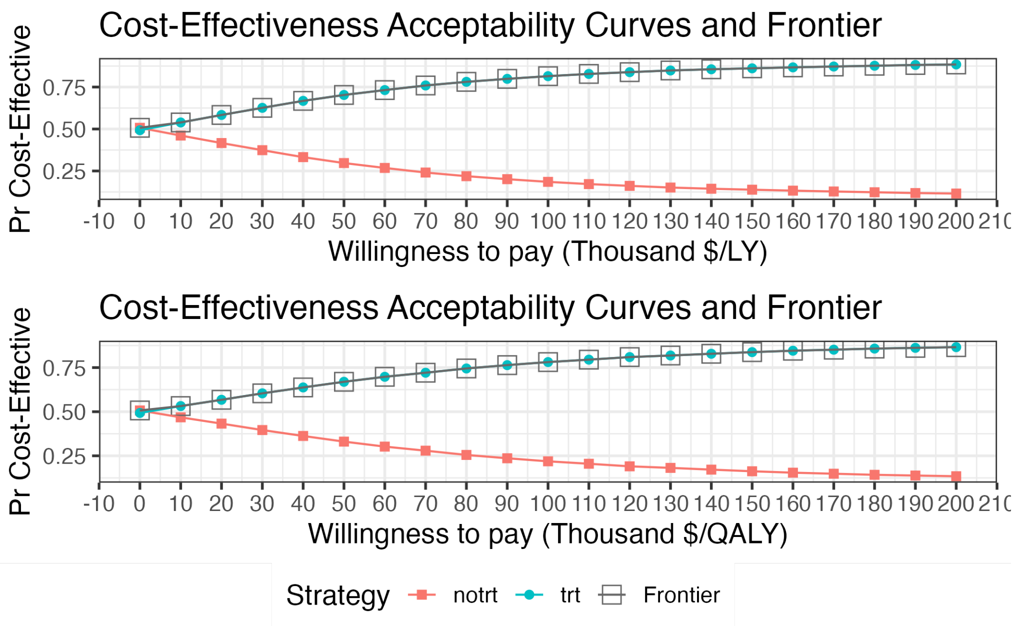

#### 3.4 EVPI and EVPPI for the population

The top panel shows the expected value of perfect (EVPI) information. The bottom panel the expected value of partial perfect (EVPPI) information.

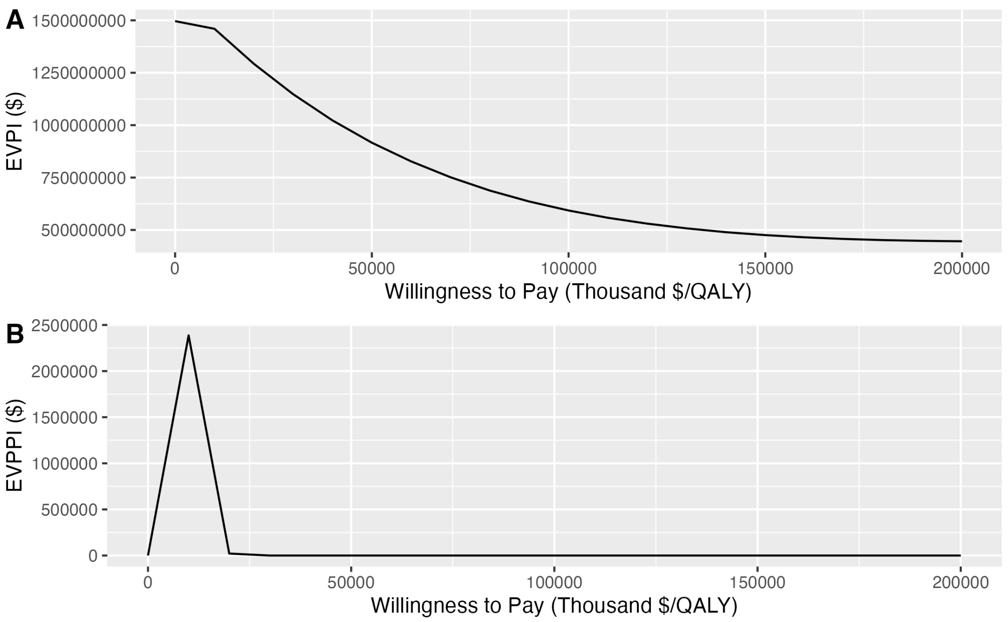

### 4 Dexamethasone

#### 4.1 CEA results

When reading the CEA results, carefully check if treatment or no treatment is shown as the reference. This has a major impact on the interpretation of the results.

| Outcome | Strategy | Cost | Effect | Inc_Cost | Inc_Effect | ICER | Status |
|---|---|---|---|---|---|---|---|
| Basecase | | | | | | | |
| QALY | notrt | 135898.2 | 4.120835 | NA | NA | NA | ND |
| QALY | trt | 142265.8 | 4.724614 | 6367.608 | 0.6037788 | 10546.260 | ND |
| PSA | | | | | | | |
| QALY | notrt | 137329.9 | 4.210067 | NA | NA | NA | ND |
| QALY | trt | 144185.7 | 4.823887 | 6855.757 | 0.6138202 | 11168.998 | ND |
| LY | notrt | 137329.9 | 6.272082 | NA | NA | NA | ND |
| LY | trt | 144185.7 | 7.194031 | 6855.757 | 0.9219491 | 7436.155 | ND |

##### 4.1.1 Uncertainty interval PA results

| | iLY | iQALY | iCosts | LY iNMB | QALY iNMB | LY iNHB | QALY iNHB |
|---|---|---|---|---|---|---|---|
| mean | 0.9219491 | 0.6138202 | 6855.757 | 85339.15 | 54526.27 | 0.8533915 | 0.5452627 |
| sd | 0.2488150 | 0.3288340 | 13652.113 | 25748.35 | 34452.59 | 0.2574835 | 0.3445259 |
| 2.5% | 0.4664479 | 0.0389291 | -19695.874 | 36840.77 | -10111.30 | 0.3680000 | -0.1010000 |
| 97.5% | 1.4342108 | 1.2584568 | 33732.899 | 137602.21 | 120436.71 | 1.3760000 | 1.2040000 |

#### 4.2 Incremental CE-plane

Incremental cost-effectiveness plane, shown as the incremental effectiveness (x-axis) and incremental cost ($) (y-axis). Iterations are represented by the small transparent dots, the mean is indicated by the large solid circle representing the results for QALY's and LY's separately. The dotted ellipses represent the 95% Credibility Intervals.

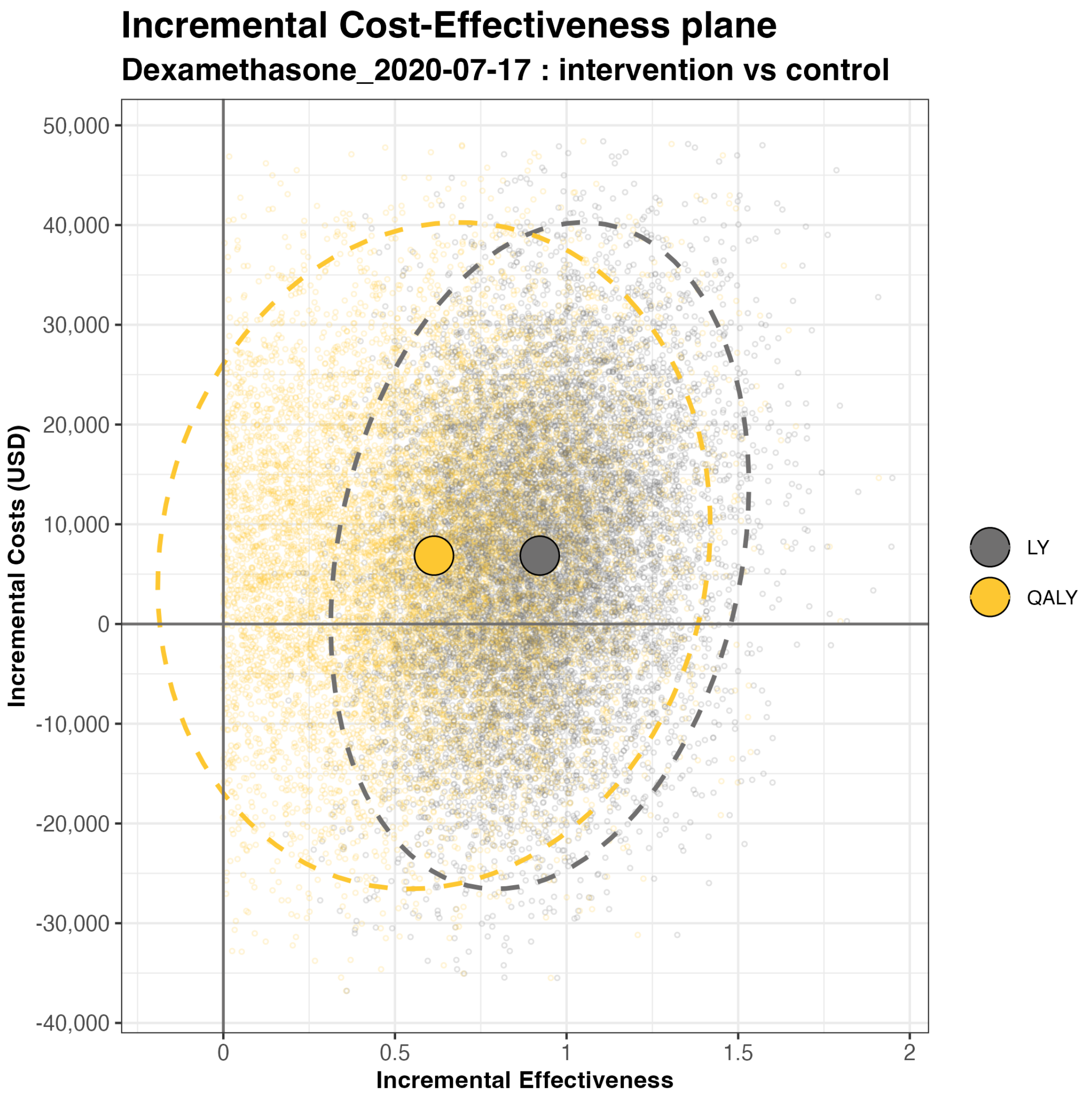
Incremental Cost-Effectiveness plane
Dexamethasone_2020-07-17 : intervention vs control
Incremental Costs (USD)
50,000
40,000
30,000
20,000
10,000
0
-10,000
-20,000
-30,000
-40,000
0
0.5
1
1.5
2
Incremental Effectiveness
LY
QALY

##### 4.3 CEAF

The figures show the cost-effectiveness acceptability curves (CEAF) with on the x-axes the willingness to pay thresholds expressed using (thousand $/QALY) in the top figure and (thousand $/LY) in the bottom figure. The y-axes represent the probability of the treatment and no treatment strategies being cost-effective.

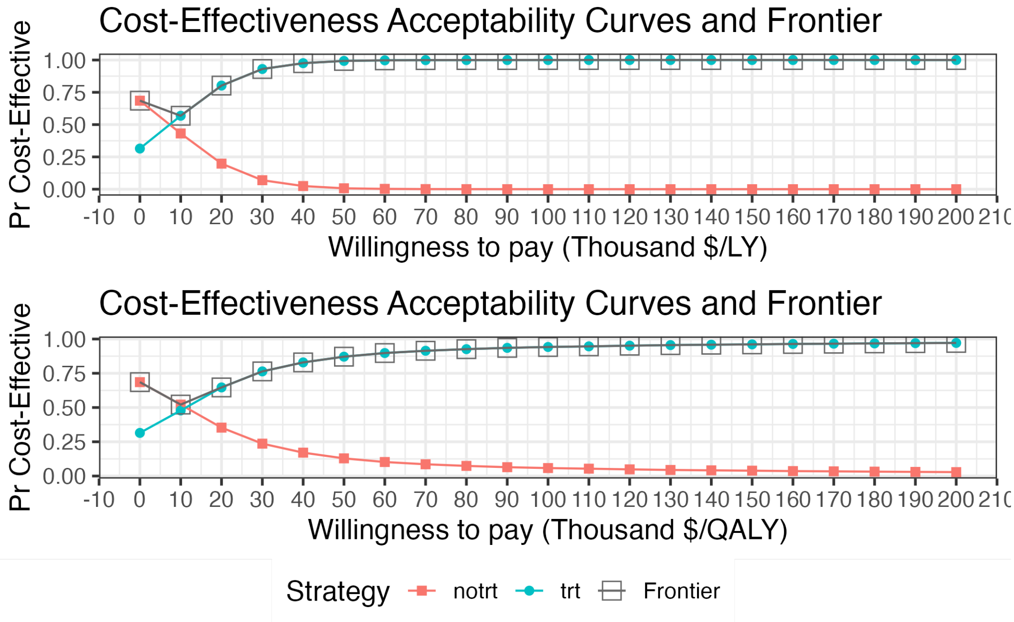

##### 4.4 EVPI and EVPPI for the population

The top panel shows the expected value of perfect (EVPI) information. The bottom panel the expected value of partial perfect (EVPPI) information.

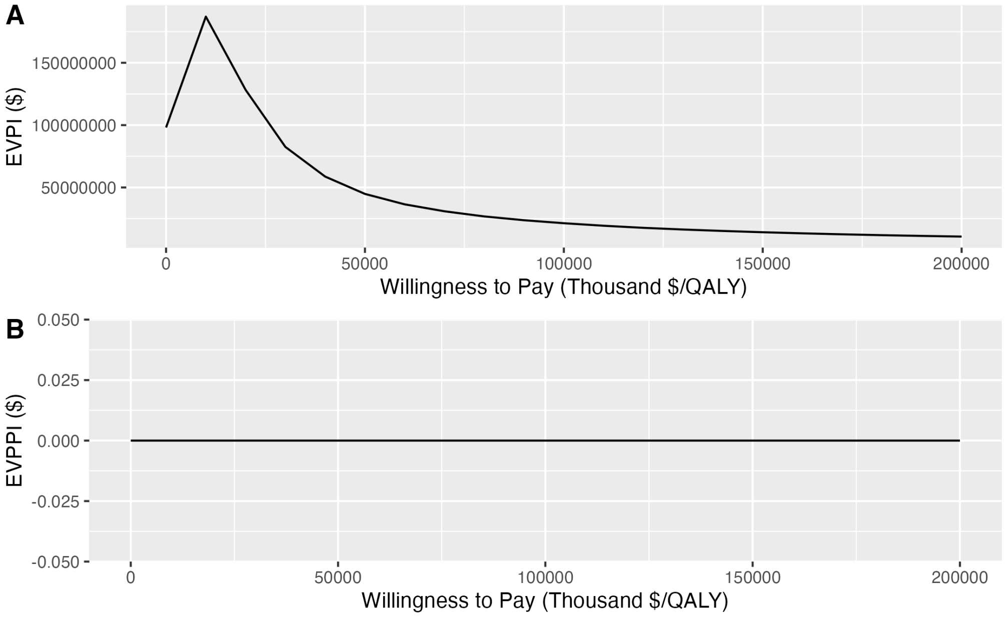

### 5 Hydroxychloroquine

#### 5.1 CEA results

When reading the CEA results, carefully check if treatment or no treatment is shown as the reference. This has a major impact on the interpretation of the results.

| Outcome | Strategy | Cost | Effect | Inc_Cost | Inc_Effect | ICER | Status |
|---|---|---|---|---|---|---|---|
| Basecase | | | | | | | |
| QALY | trt | 119253.0 | 5.809026 | NA | NA | NA | ND |
| QALY | notrt | 131616.0 | 6.060284 | 12362.96 | 0.2512585 | 49204.15 | ND |
| PSA | | | | | | | |
| QALY | trt | 120317.8 | 5.952873 | NA | NA | NA | ND |
| QALY | notrt | 132544.5 | 6.216225 | 12226.73 | 0.2633519 | 46427.35 | ND |
| LY | trt | 120317.8 | 7.003557 | NA | NA | NA | ND |
| LY | notrt | 132544.5 | 7.326196 | 12226.73 | 0.3226389 | 37896.02 | ND |

##### 5.1.1 Uncertainty interval PA results

| | iLY | iQALY | iCosts | LY iNMB | QALY iNMB | LY iNHB | QALY iNHB |
|---|---|---|---|---|---|---|---|
| mean | -0.3226389 | -0.2633519 | -12226.728 | -20037.16 | -14108.46 | -0.2003716 | -0.1410846 |
| sd | 0.2477098 | 0.2063808 | 10310.542 | 23810.57 | 20305.45 | 0.2381057 | 0.2030545 |
| 2.5% | -0.8533022 | -0.7125399 | -32724.776 | -70511.85 | -57220.76 | -0.7050000 | -0.5720000 |
| 97.5% | 0.1170741 | 0.0983651 | 7344.807 | 22945.99 | 22481.89 | 0.2290000 | 0.2250000 |

#### 5.2 Incremental CE-plane

Incremental cost-effectiveness plane, shown as the incremental effectiveness (x-axis) and incremental cost ($) (y-axis). Iterations are represented by the small transparent dots, the mean is indicated by the large solid circle representing the results for QALY's and LY's separately. The dotted ellipses represent the 95% Credibility Intervals.

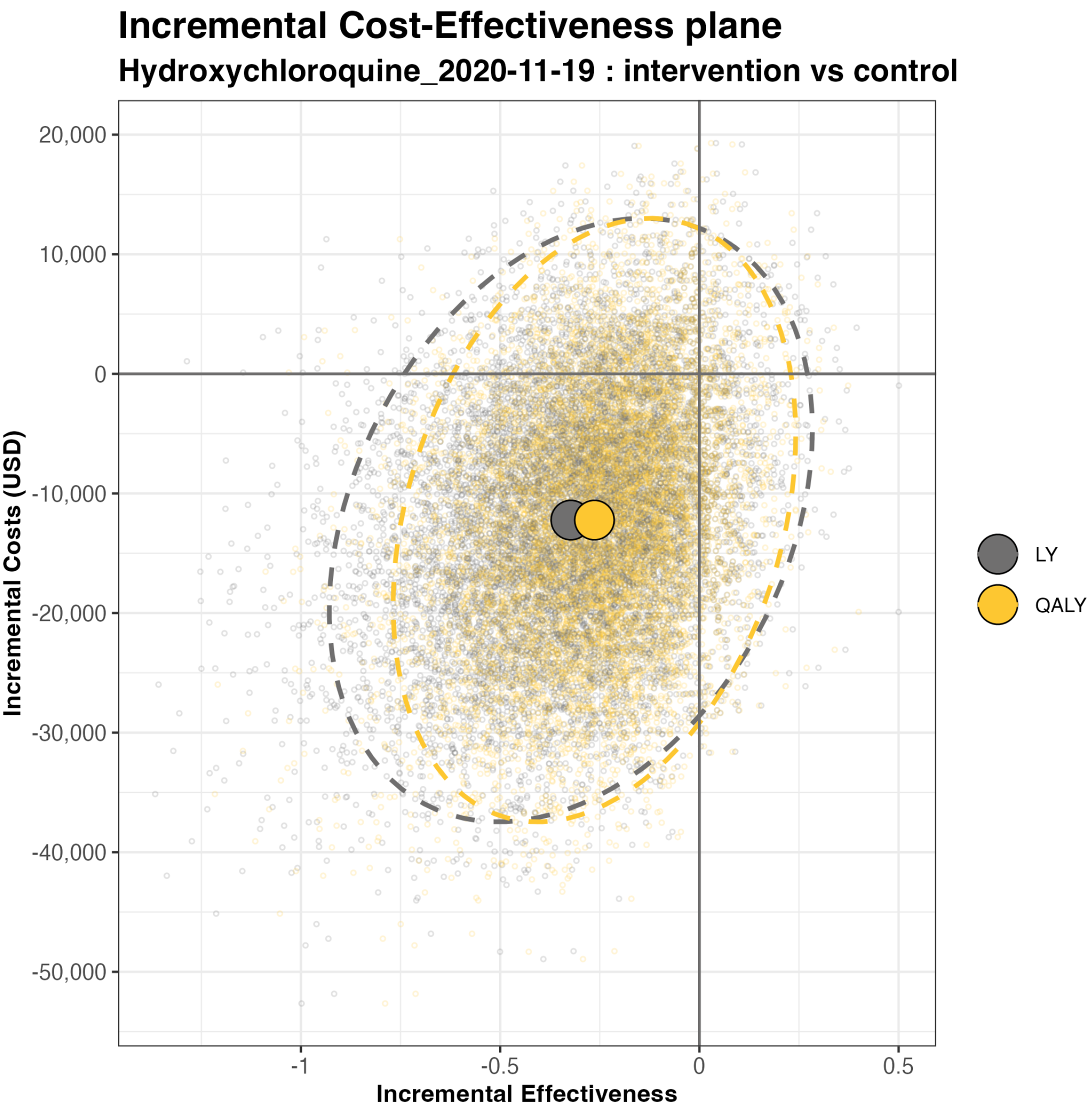

Incremental Cost-Effectiveness plane
Hydroxychloroquine_2020-11-19 : intervention vs control
20,000
10,000
0
-10,000
-20,000
-30,000
-40,000
-50,000
Incremental Costs (USD)
-1
-0.5
0
0.5
Incremental Effectiveness
LY
QALY

#### 5.3 CEAF

The figures show the cost-effectiveness acceptability curves (CEAF) with on the x-axes the willingness to pay thresholds expressed using (thousand $/QALY) in the top figure and (thousand $/LY) in the bottom figure. The y-axes represent the probability of the treatment and no treatment strategies being cost-effective.

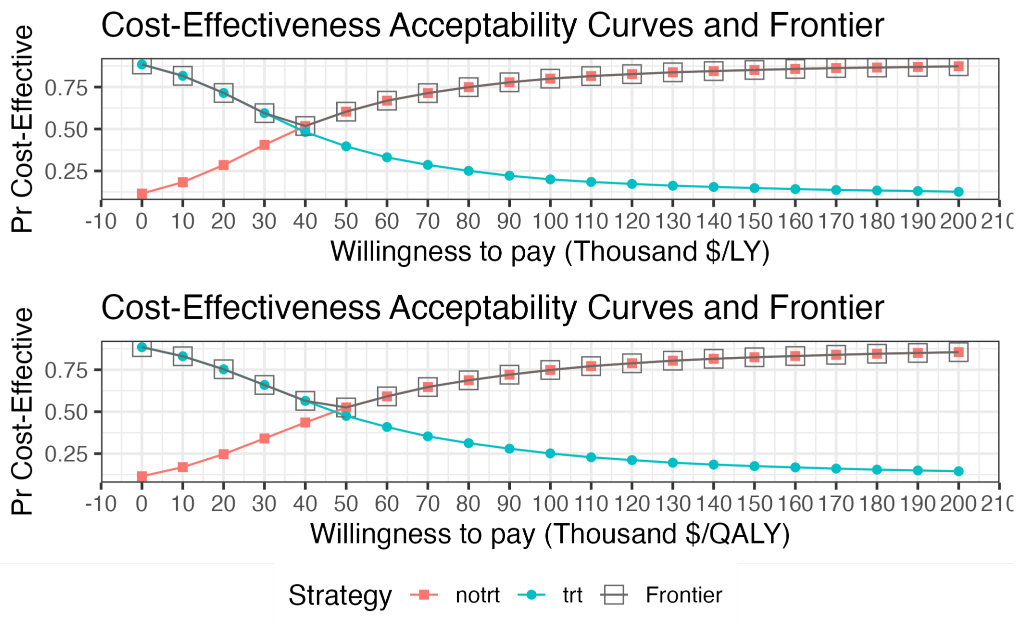

#### 5.4 EVPI and EVPPI for the population

The top panel shows the expected value of perfect (EVPI) information. The bottom panel the expected value of partial perfect (EVPPI) information.

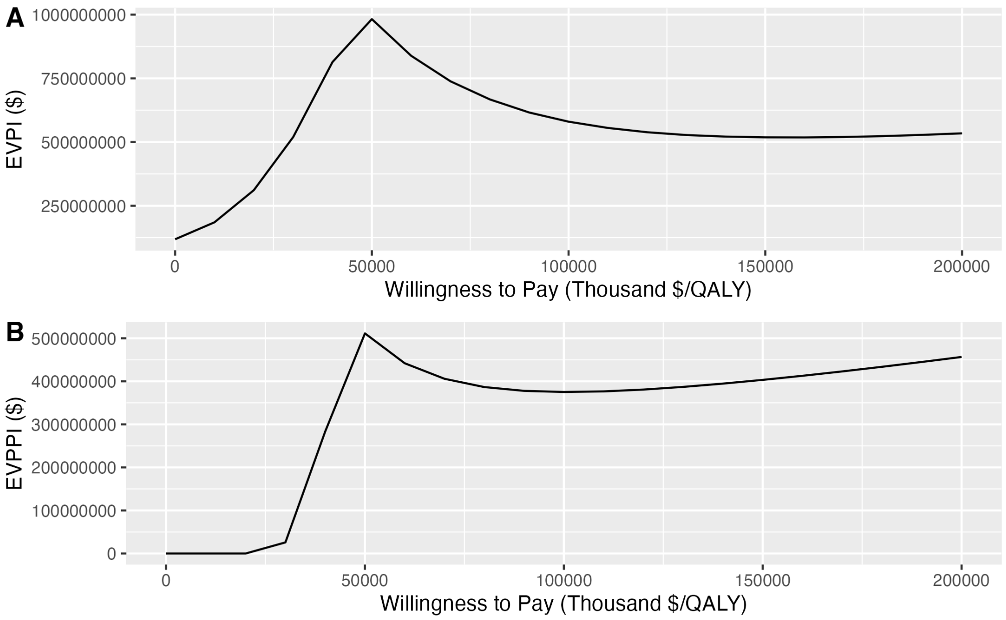

#### 5.5 Expected net benefit

The expected net benefit for Hydroxychloroquine. The x-axis gives the study sample size, and the y-axis corresponding expected net benefit.

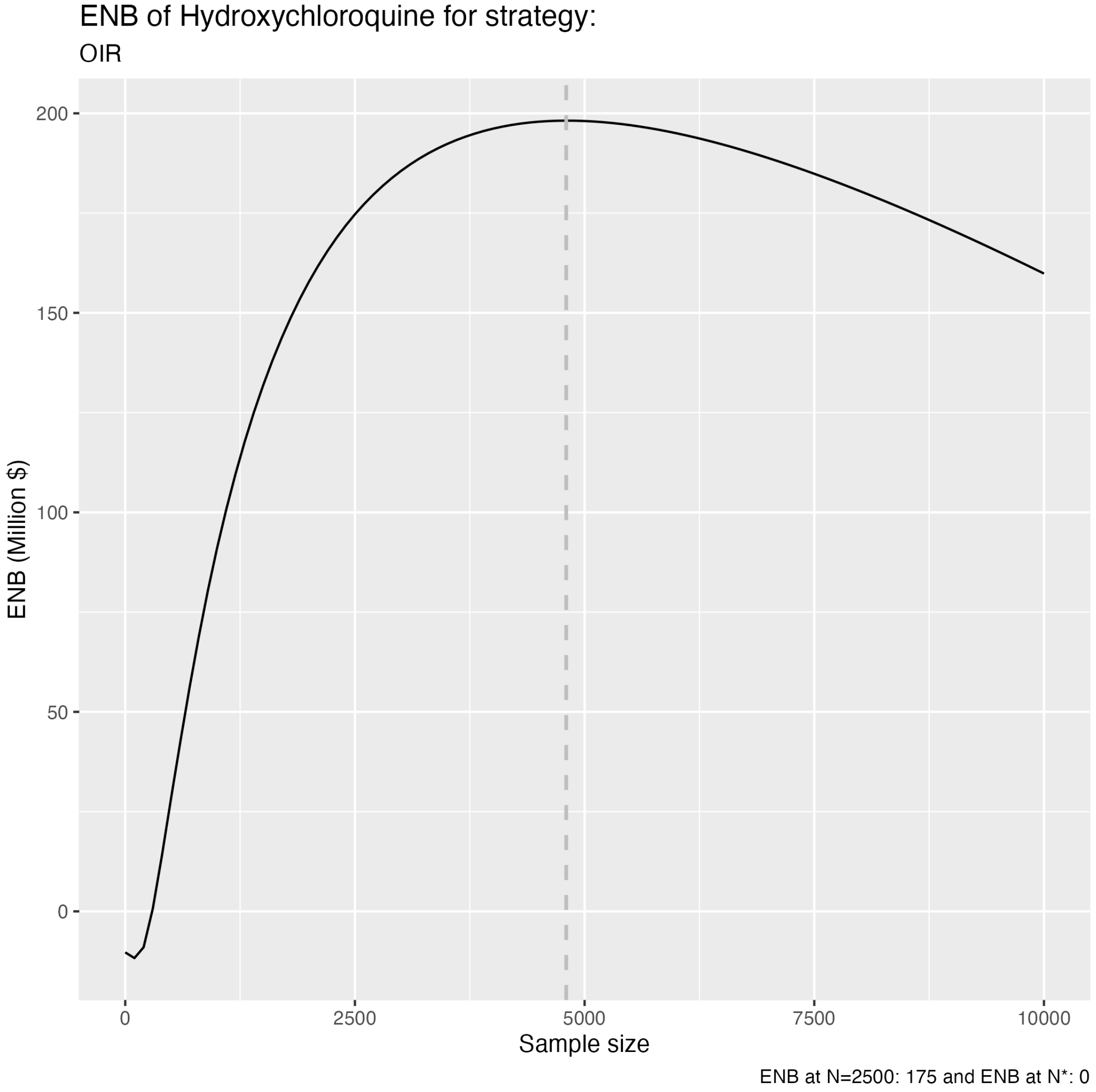

### 6 Interferon-B1a

#### 6.1 CEA results

When reading the CEA results, carefully check if treatment or no treatment is shown as the reference. This has a major impact on the interpretation of the results.

| Outcome | Strategy | Cost | Effect | Inc_Cost | Inc_Effect | ICER | Status |
|---|---|---|---|---|---|---|---|
| Basecase | | | | | | | |
| QALY | trt | 100332.7 | 5.615119 | NA | NA | NA | ND |
| QALY | notrt | 102493.9 | 6.063533 | 2161.158 | 0.4484143 | 4819.555 | ND |
| PSA | | | | | | | |
| QALY | trt | 101612.5 | 5.746150 | NA | NA | NA | ND |
| QALY | notrt | 104150.7 | 6.218156 | 2538.203 | 0.4720061 | 5377.478 | ND |
| LY | trt | 101612.5 | 6.743815 | NA | NA | NA | ND |
| LY | notrt | 104150.7 | 7.316302 | 2538.203 | 0.5724873 | 4433.640 | ND |

##### 6.1.1 Uncertainty interval PA results

| | iLY | iQALY | iCosts | LY iNMB | QALY iNMB | LY iNHB | QALY iNHB |
|---|---|---|---|---|---|---|---|
| mean | -0.5724873 | -0.4720061 | -2538.203 | -54710.523 | -44662.409 | -0.5471052 | -0.4466241 |
| sd | 0.3980288 | 0.3311650 | 5852.043 | 35236.833 | 28648.811 | 0.3523683 | 0.2864881 |
| 2.5% | -1.4321205 | -1.1898927 | -14453.172 | -130606.536 | -106088.419 | -1.3060000 | -1.0610000 |
| 97.5% | 0.1208941 | 0.0998084 | 8622.124 | 7595.372 | 5378.487 | 0.0760000 | 0.0540000 |

#### 6.2 Incremental CE-plane

Incremental cost-effectiveness plane, shown as the incremental effectiveness (x-axis) and incremental cost ($) (y-axis). Iterations are represented by the small transparent dots, the mean is indicated by the large solid circle representing the results for QALY's and LY's separately. The dotted ellipses represent the 95% Credibility Intervals.

#### Incremental Cost-Effectiveness plane

##### Interferon-B1a_2020-12-02 : intervention vs control

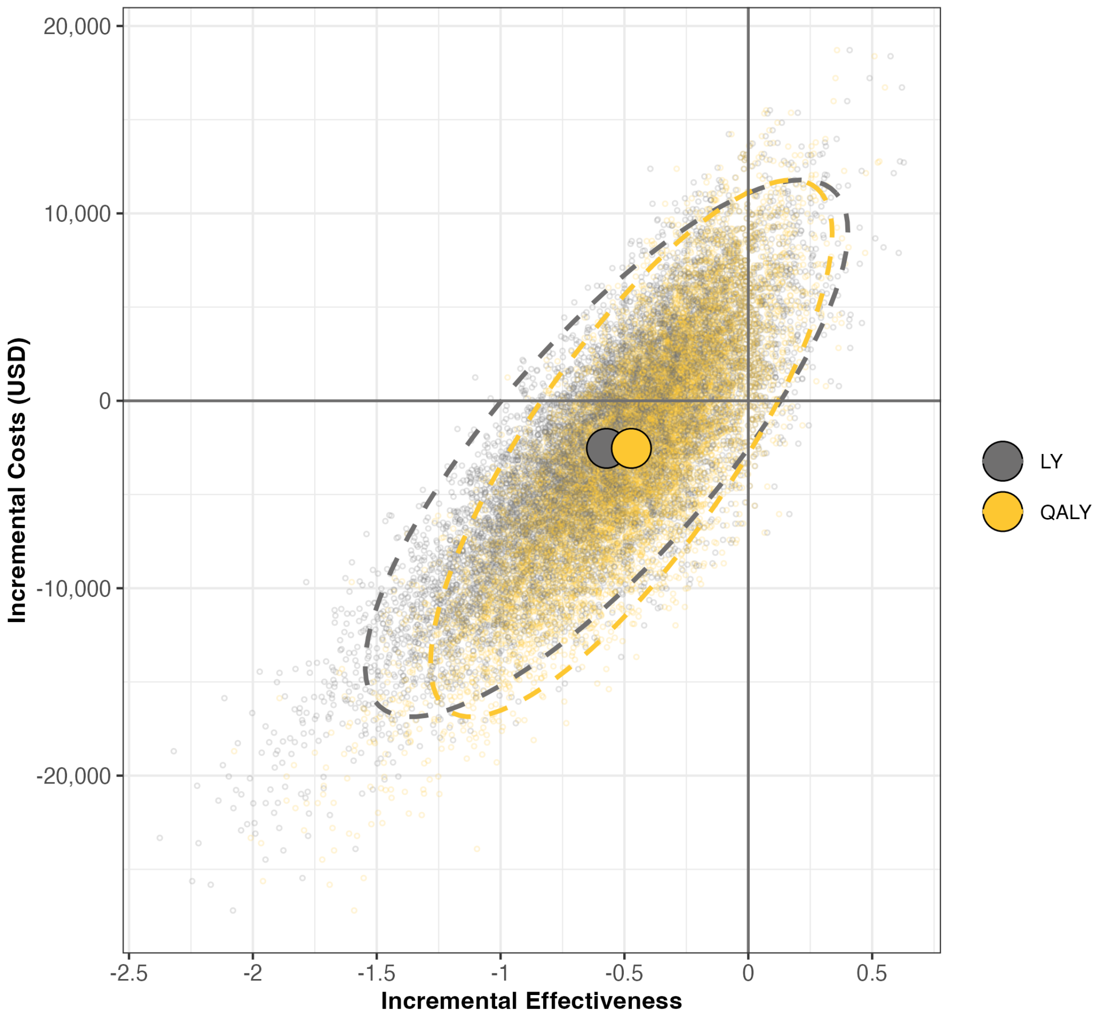

#### 6.3 CEAF

The figures show the cost-effectiveness acceptability curves (CEAF) with on the x-axes the willingness to pay thresholds expressed using (thousand $/QALY) in the top figure and (thousand $/LY) in the bottom figure. The y-axes represent the probability of the treatment and no treatment strategies being cost-effective.

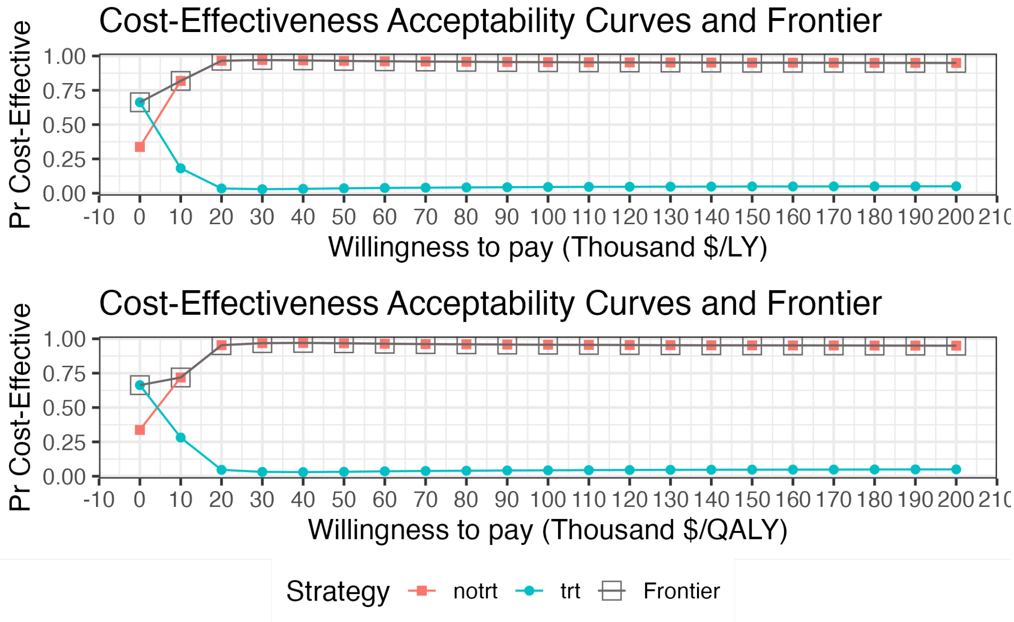

#### 6.4 EVPI and EVPPI for the population

The top panel shows the expected value of perfect (EVPI) information. The bottom panel the expected value of partial perfect (EVPPI) information.

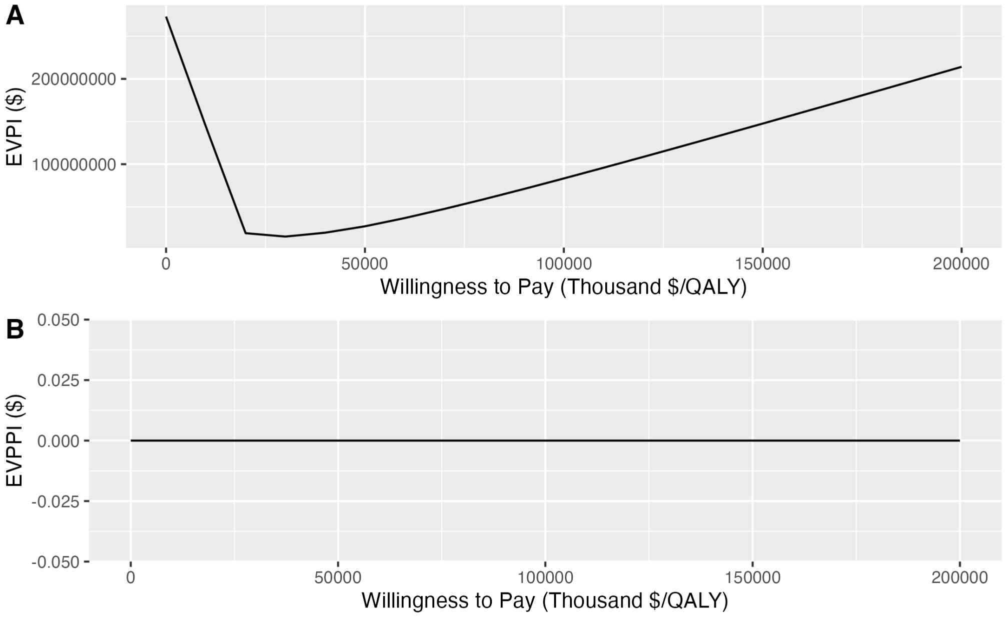

### 7 Lopinavir-Ritonavir

#### 7.1 CEA results

When reading the CEA results, carefully check if treatment or no treatment is shown as the reference. This has a major impact on the interpretation of the results.

| Outcome | Strategy | Cost | Effect | Inc_Cost | Inc_Effect | ICER | Status |
|---|---|---|---|---|---|---|---|
| Basecase | | | | | | | |
| QALY | trt | 115845.9 | 5.977831 | NA | NA | NA | ND |
| QALY | notrt | 117054.9 | 6.061909 | 1208.970 | 0.0840777 | 14379.20 | ND |
| PSA | | | | | | | |
| QALY | trt | 127344.3 | 6.125459 | NA | NA | NA | ND |
| QALY | notrt | 128748.2 | 6.216511 | 1403.841 | 0.0910525 | 15417.93 | ND |
| LY | trt | 127344.3 | 7.214606 | NA | NA | NA | ND |
| LY | notrt | 128748.2 | 7.324898 | 1403.841 | 0.1102931 | 12728.28 | ND |

##### 7.1.1 Uncertainty interval PA results

| | iLY | iQALY | iCosts | LY iNMB | QALY iNMB | LY iNHB | QALY iNHB |
|---|---|---|---|---|---|---|---|
| mean | -0.1102931 | -0.0910525 | -1403.841 | -9625.465 | -7701.405 | -0.0962547 | -0.0770140 |
| sd | 0.2378630 | 0.1972737 | 19815.084 | 28235.828 | 26003.919 | 0.2823583 | 0.2600392 |
| 2.5% | -0.6064263 | -0.5011920 | -40583.542 | -67312.440 | -59992.078 | -0.6730000 | -0.6000000 |
| 97.5% | 0.3376302 | 0.2763129 | 37062.206 | 44844.583 | 42960.653 | 0.4480000 | 0.4300000 |

#### 7.2 Incremental CE-plane

Incremental cost-effectiveness plane, shown as the incremental effectiveness (x-axis) and incremental cost ($) (y-axis). Iterations are represented by the small transparent dots, the mean is indicated by the large solid circle representing the results for QALY's and LY's separately. The dotted ellipses represent the 95% Credibility Intervals.

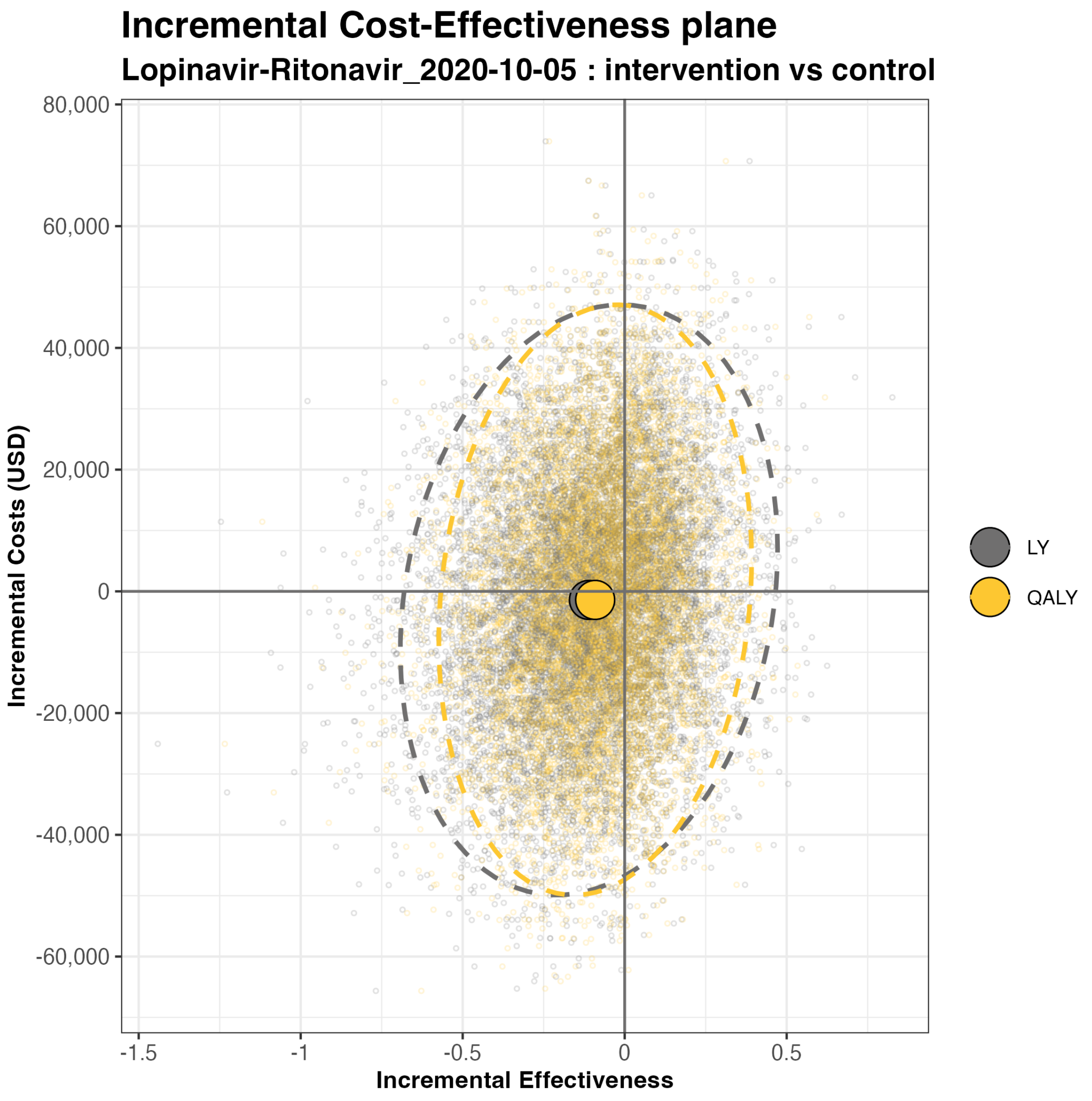
Incremental Cost-Effectiveness plane
Lopinavir-Ritonavir_2020-10-05 : intervention vs control
Incremental Costs (USD)
80,000
60,000
40,000
20,000
0
-20,000
-40,000
-60,000
-1.5
-1
-0.5
0
0.5
Incremental Effectiveness
LY
QALY

#### 7.3 CEAF

The figures show the cost-effectiveness acceptability curves (CEAF) with on the x-axes the willingness to pay thresholds expressed using (thousand $/QALY) in the top figure and (thousand $/LY) in the bottom figure. The y-axes represent the probability of the treatment and no treatment strategies being cost-effective.

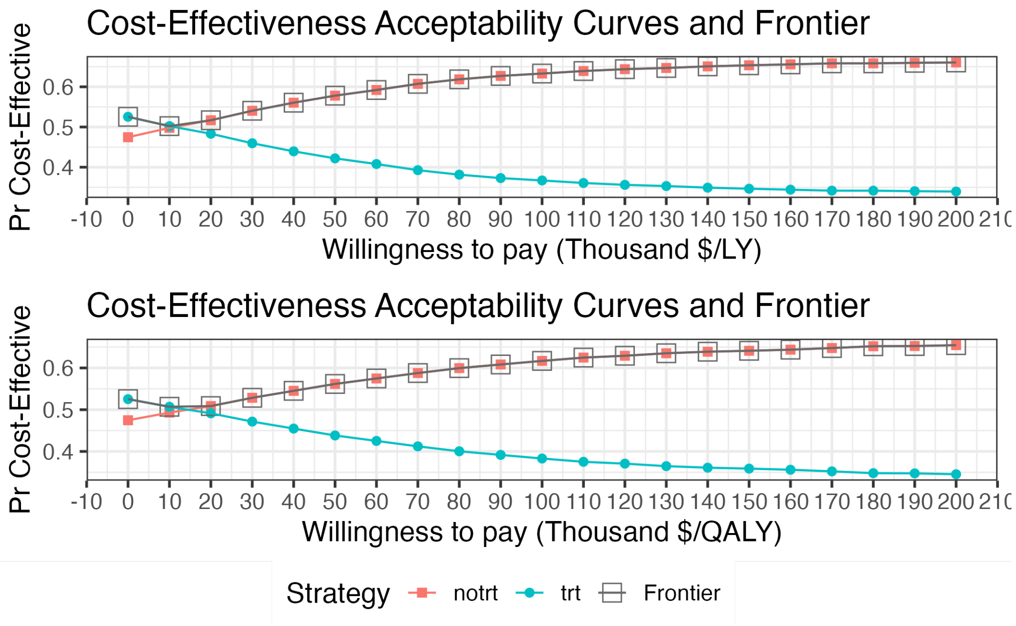

#### 7.4 EVPI and EVPPI for the population

The top panel shows the expected value of perfect (EVPI) information. The bottom panel the expected value of partial perfect (EVPPI) information.

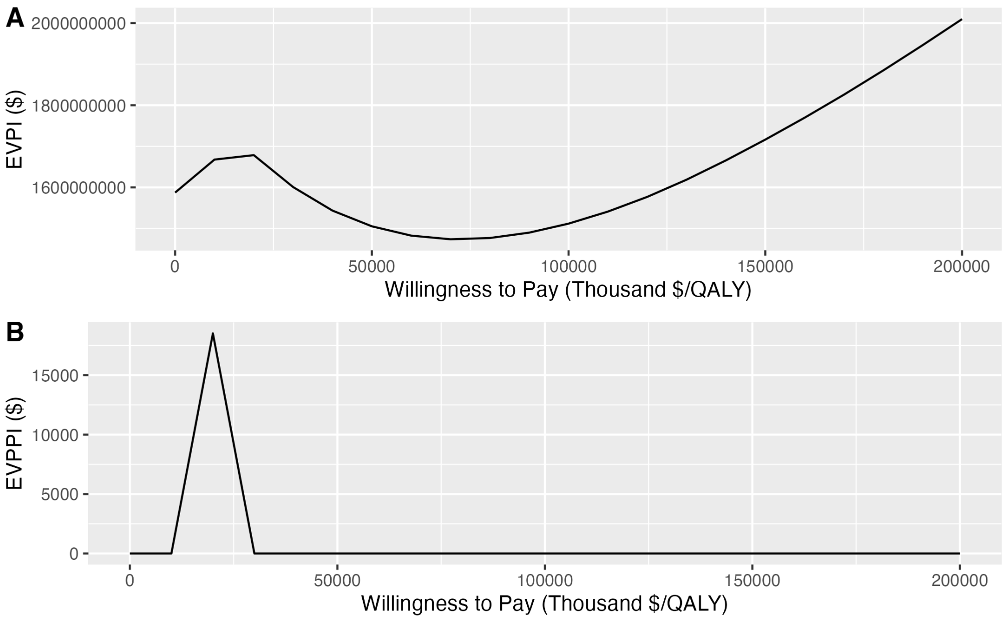

### 8 Remdesivir

#### 8.1 CEA results

When reading the CEA results, carefully check if treatment or no treatment is shown as the reference. This has a major impact on the interpretation of the results.

| Outcome | Strategy | Cost | Effect | Inc_Cost | Inc_Effect | ICER | Status |
|---|---|---|---|---|---|---|---|
| Basecase | | | | | | | |
| QALY | trt | 126468.2 | 6.313817 | NA | NA | NA | ND |
| QALY | notrt | 134528.2 | 6.059959 | NA | NA | NA | D |
| PSA | | | | | | | |
| QALY | trt | 137220.4 | 6.468324 | NA | NA | NA | ND |
| QALY | notrt | 137225.9 | 6.215891 | NA | NA | NA | D |
| LY | trt | 137220.4 | 7.630518 | NA | NA | NA | ND |
| LY | notrt | 137225.9 | 7.327818 | NA | NA | NA | D |

##### 8.1.1 Uncertainty interval PA results

| | iLY | iQALY | iCosts | LY iNMB | QALY iNMB | LY iNHB | QALY iNHB |
|---|---|---|---|---|---|---|---|
| mean | 0.3027006 | 0.2524324 | -5.49902 | 30275.56 | 25248.74 | 0.3027556 | 0.2524874 |
| sd | 0.2443364 | 0.2037589 | 17736.94729 | 27327.87 | 24789.38 | 0.2732787 | 0.2478938 |
| 2.5% | -0.1759477 | -0.1406231 | -33318.48680 | -23624.02 | -23881.16 | -0.2360000 | -0.2390000 |
| 97.5% | 0.7868397 | 0.6702243 | 35724.00724 | 83113.69 | 73205.91 | 0.8310000 | 0.7320000 |

#### 8.2 Incremental CE-plane

Incremental cost-effectiveness plane, shown as the incremental effectiveness (x-axis) and incremental cost ($) (y-axis). Iterations are represented by the small transparent dots, the mean is indicated by the large solid circle representing the results for QALY's and LY's separately. The dotted ellipses represent the 95% Credibility Intervals.

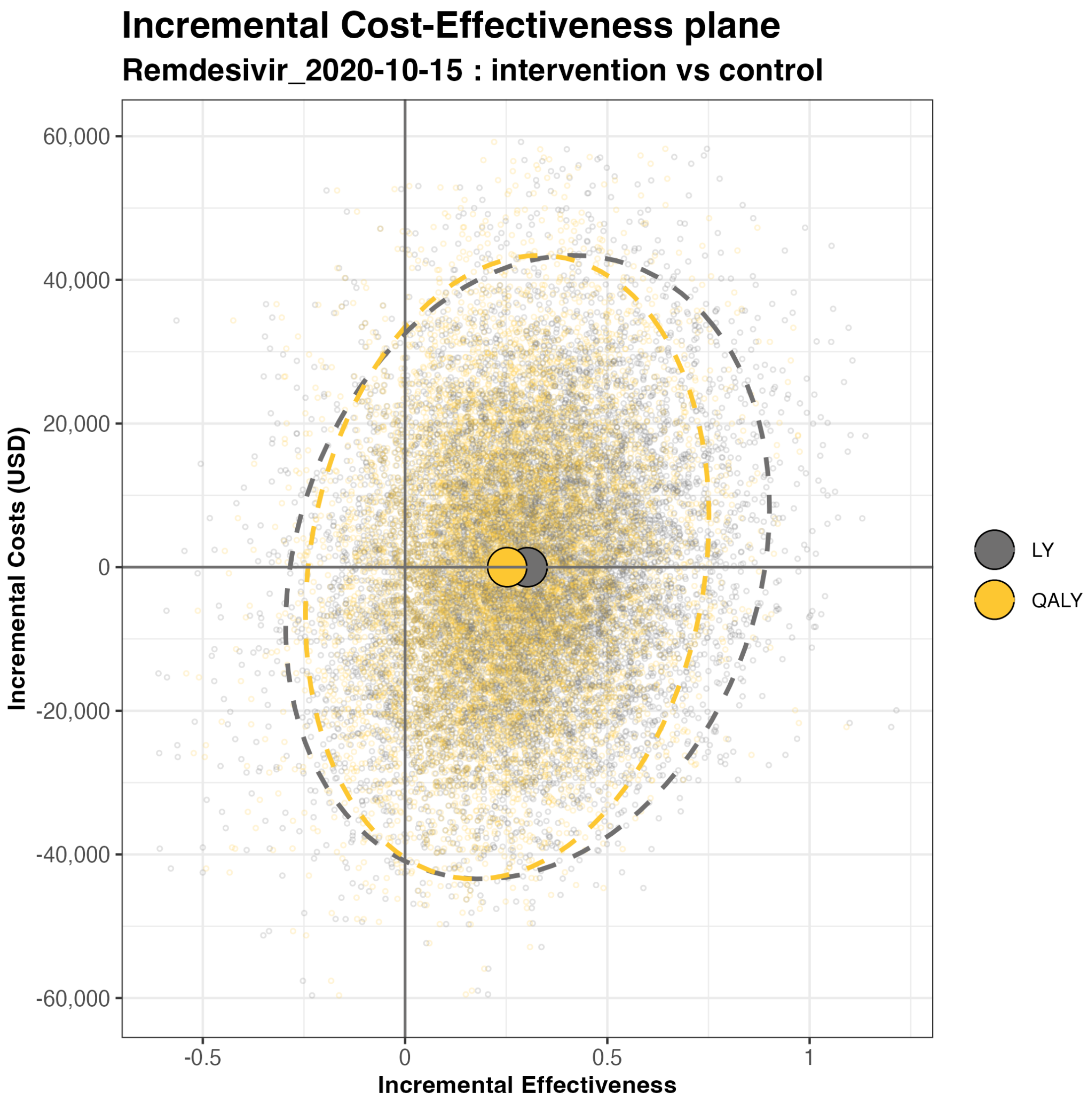

Incremental Cost-Effectiveness plane
Remdesivir_2020-10-15 : intervention vs control
Incremental Costs (USD)
60,000
40,000
20,000
0
-20,000
-40,000
-60,000
-0.5
0
0.5
1
Incremental Effectiveness
LY
QALY

#### 8.3 CEAF

The figures show the cost-effectiveness acceptability curves (CEAF) with on the x-axes the willingness to pay thresholds expressed using (thousand $/QALY) in the top figure and (thousand $/LY) in the bottom figure. The y-axes represent the probability of the treatment and no treatment strategies being cost-effective.

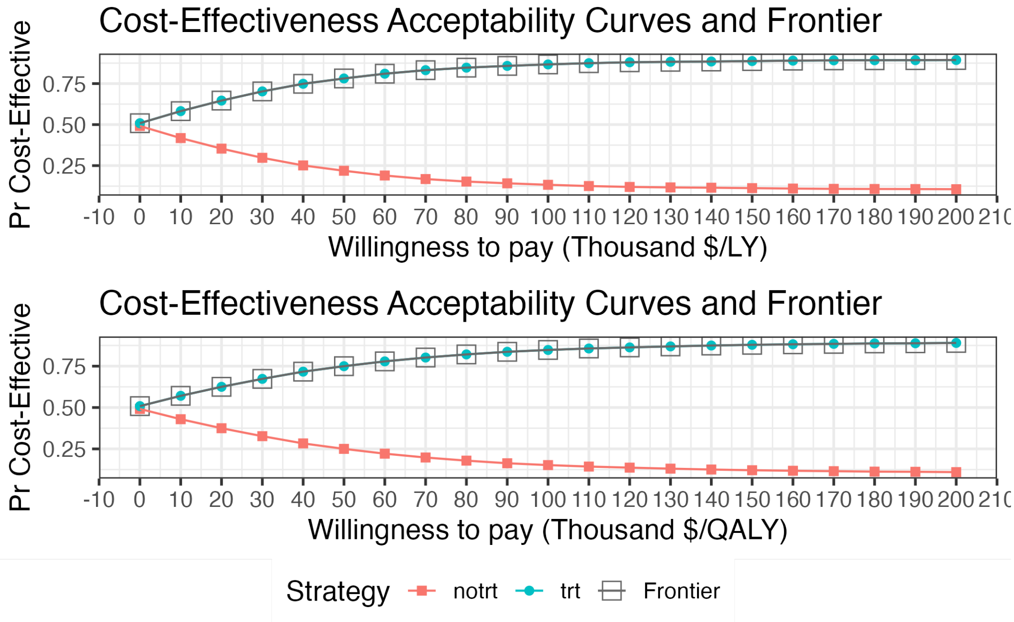

#### 8.4 EVPI and EVPPI for the population

The top panel shows the expected value of perfect (EVPI) information. The bottom panel the expected value of partial perfect (EVPPI) information.

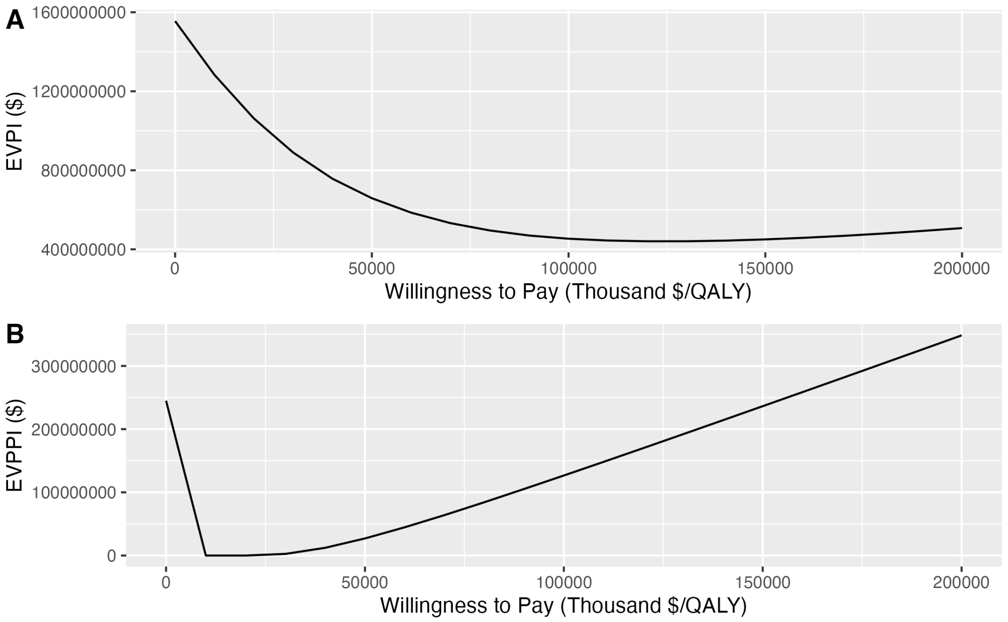

### 9 Tocilizumab

#### 9.1 CEA results

When reading the CEA results, carefully check if treatment or no treatment is shown as the reference. This has a major impact on the interpretation of the results.

| Outcome | Strategy | Cost | Effect | Inc_Cost | Inc_Effect | ICER | Status |
|---|---|---|---|---|---|---|---|
| Basecase | | | | | | | |
| QALY | notrt | 110381.3 | 4.150210 | NA | NA | NA | ND |
| QALY | trt | 145945.4 | 5.018784 | 35564.11 | 0.8685746 | 40945.38 | ND |
| PSA | | | | | | | |
| QALY | notrt | 113659.5 | 4.243665 | NA | NA | NA | ND |
| QALY | trt | 149509.0 | 5.125943 | 35849.42 | 0.8822780 | 40632.79 | ND |
| LY | notrt | 113659.5 | 6.311468 | NA | NA | NA | ND |
| LY | trt | 149509.0 | 7.637100 | 35849.42 | 1.3256321 | 27043.27 | ND |

##### 9.1.1 Uncertainty interval PA results

| | iLY | iQALY | iCosts | LY iNMB | QALY iNMB | LY iNHB | QALY iNHB |
|---|---|---|---|---|---|---|---|
| mean | 1.3256321 | 0.8822780 | 35849.422 | 96713.79 | 52378.38 | 0.9671379 | 0.5237838 |
| sd | 0.4275917 | 0.5039017 | 8081.919 | 38079.88 | 47964.87 | 0.3807988 | 0.4796487 |
| 2.5% | 0.5435232 | 0.0518686 | 20447.944 | 27669.74 | -30048.71 | 0.2770000 | -0.3000000 |
| 97.5% | 2.2190714 | 1.9372177 | 52175.482 | 175794.14 | 149555.26 | 1.7580000 | 1.4960000 |

#### 9.2 Incremental CE-plane

Incremental cost-effectiveness plane, shown as the incremental effectiveness (x-axis) and incremental cost ($) (y-axis). Iterations are represented by the small transparent dots, the mean is indicated by the large solid circle representing the results for QALY's and LY's separately. The dotted ellipses represent the 95% Credibility Intervals.

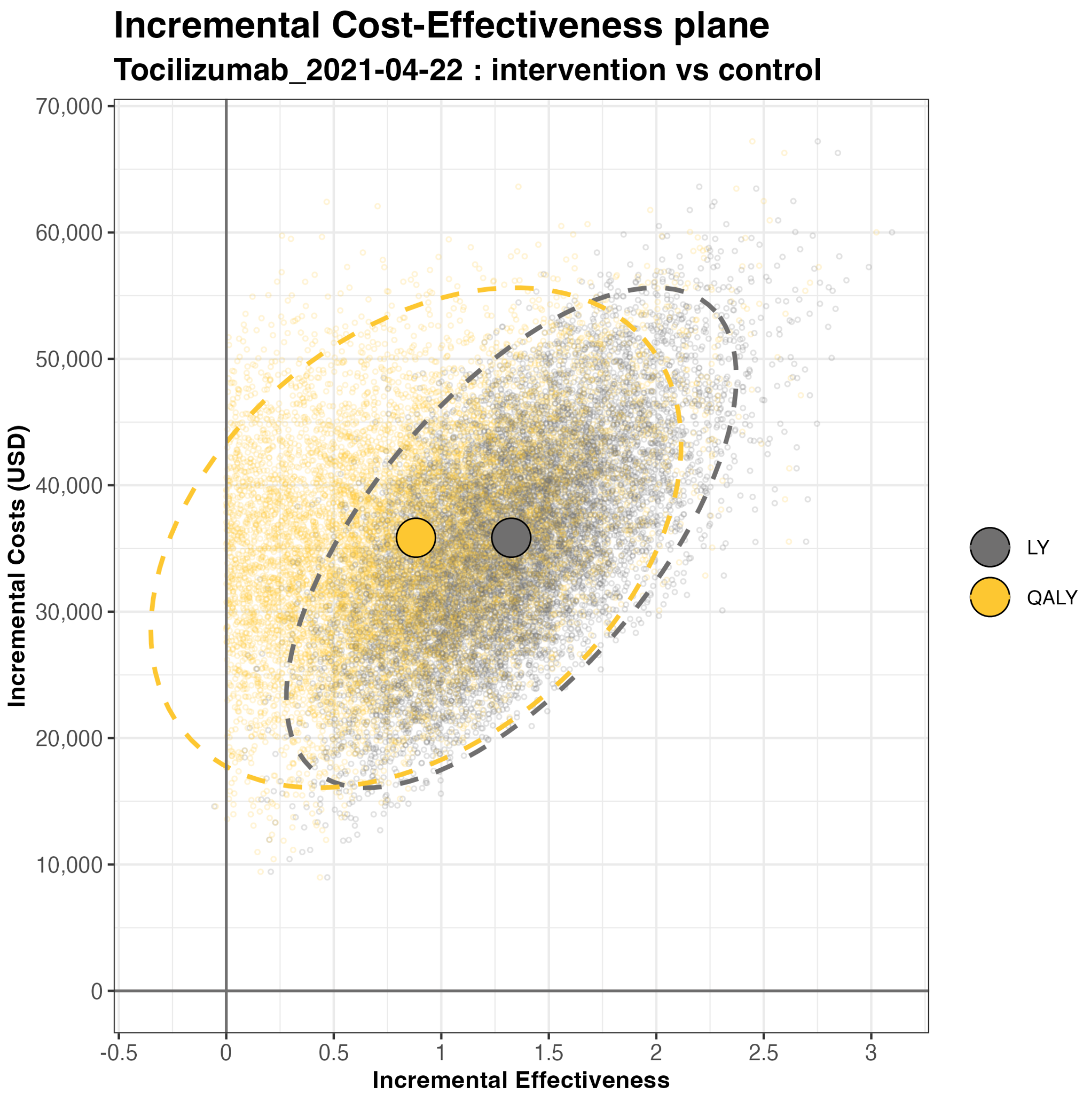

Incremental Cost-Effectiveness plane
Tocilizumab_2021-04-22 : intervention vs control
70,000
60,000
50,000
40,000
30,000
20,000
10,000
0
Incremental Costs (USD)
-0.5
0
0.5
1
1.5
2
2.5
3
Incremental Effectiveness
LY
QALY

#### 9.3 CEAF

The figures show the cost-effectiveness acceptability curves (CEAF) with on the x-axes the willingness to pay thresholds expressed using (thousand $/QALY) in the top figure and (thousand $/LY) in the bottom figure. The y-axes represent the probability of the treatment and no treatment strategies being cost-effective.

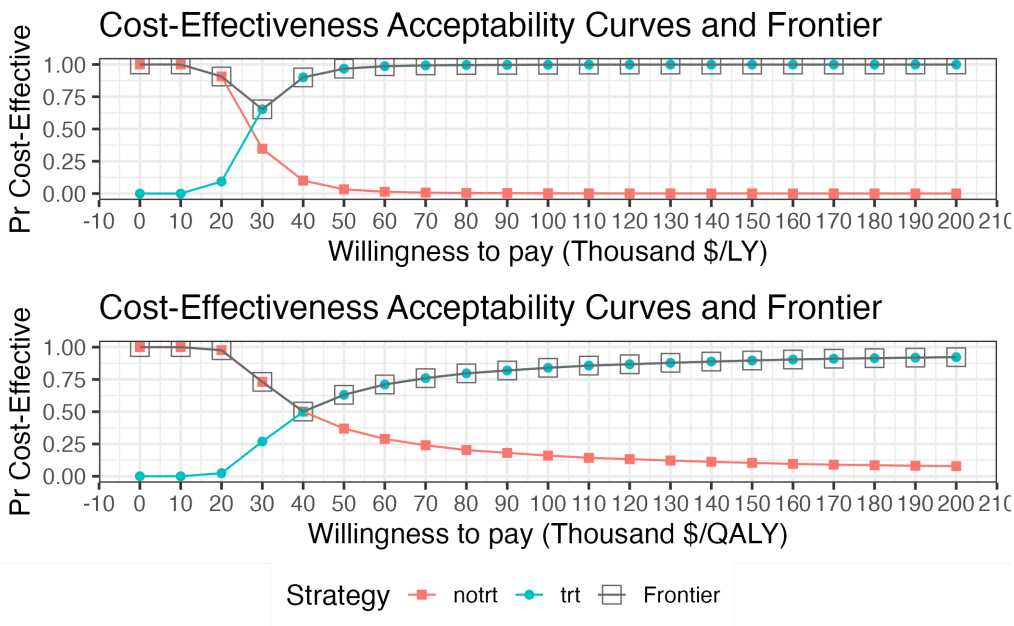

#### 9.4 EVPI and EVPPI for the population

The top panel shows the expected value of perfect (EVPI) information. The bottom panel the expected value of partial perfect (EVPPI) information.

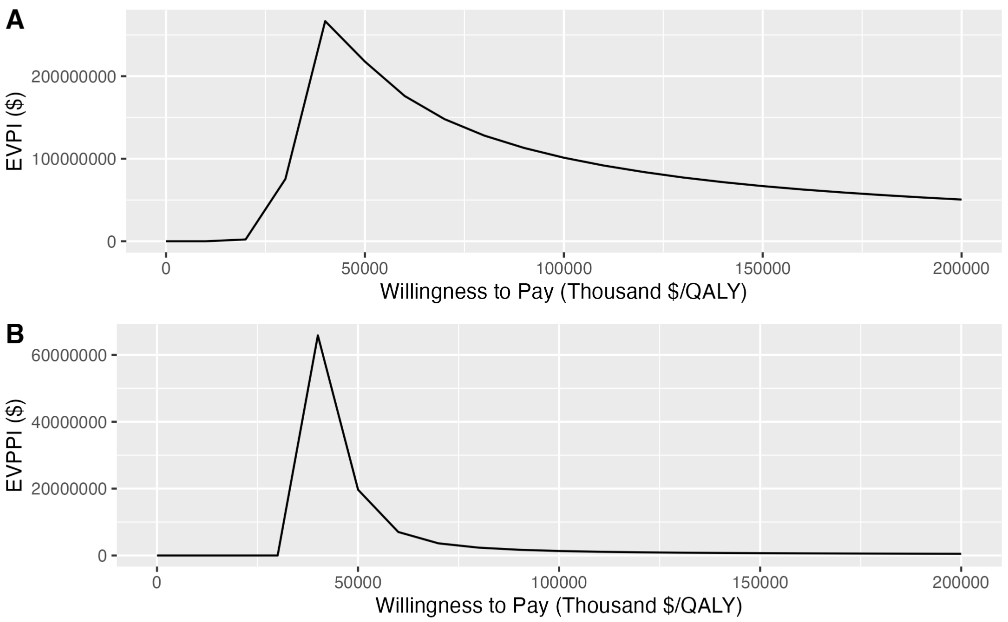

### Probabilistic analysis – parameter distributions

#### Cohort distributions

This are the distributions of the input parameters used in the probabilistic analysis. These input parameters are the same for all treatments.

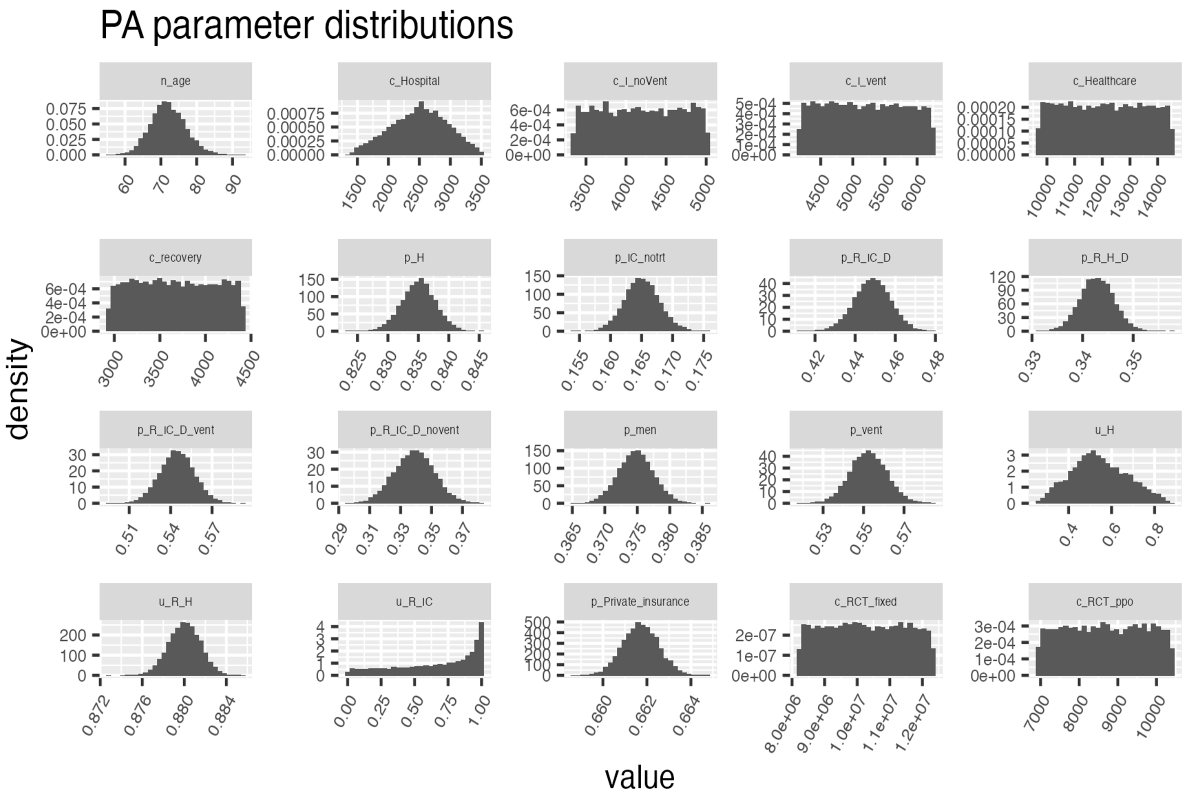

#### Treatment specific distributions

This are the distributions that are used for the different treatments

#### Baricitinib-Remdesivir

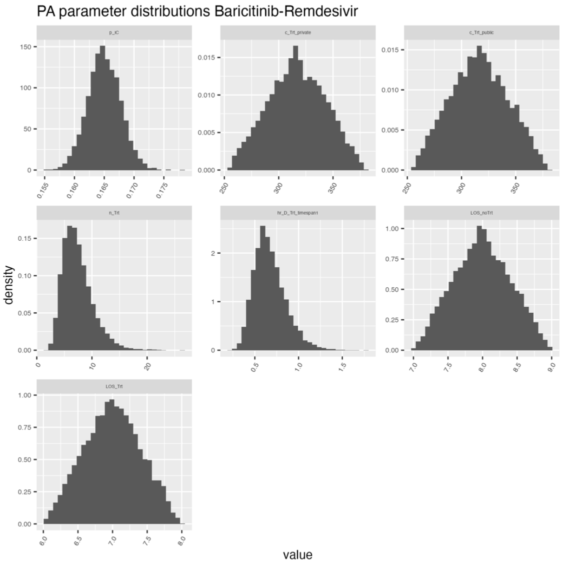

#### Casirivimab-Imdevimab

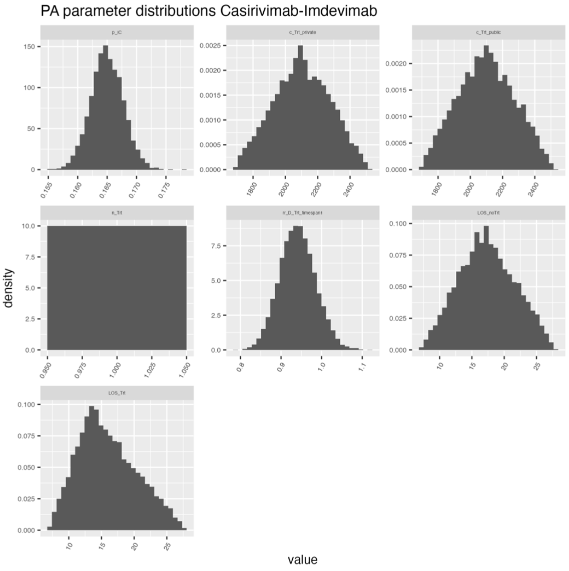

### Dexamethasone

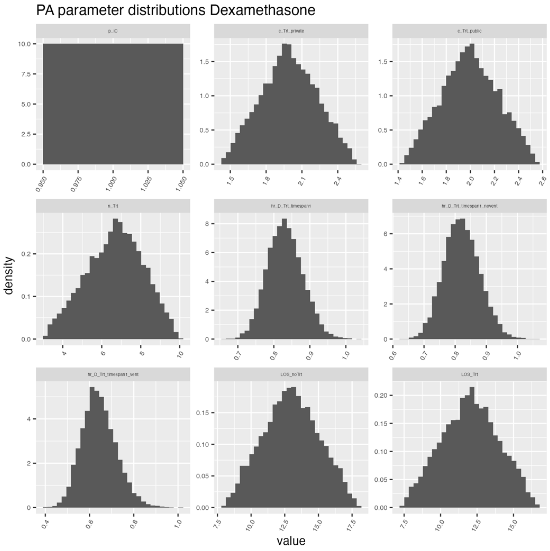

### Hydroxychloroquine

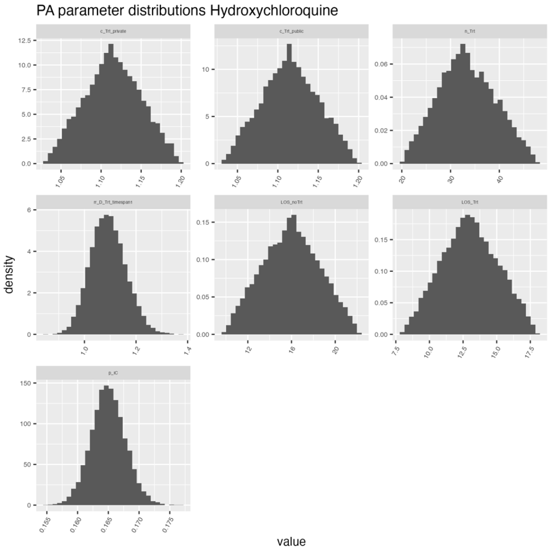

### Interferon-B1a

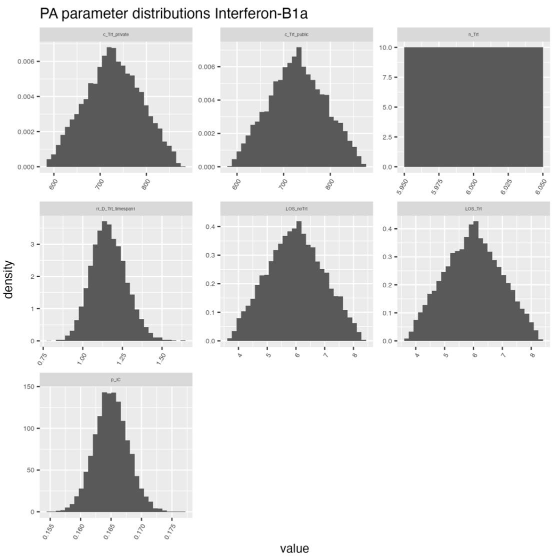

### Lopinavir-Ritonavir

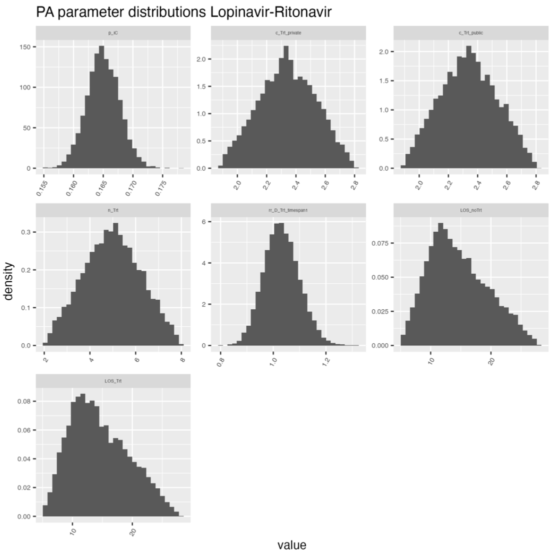

### Remdesivir

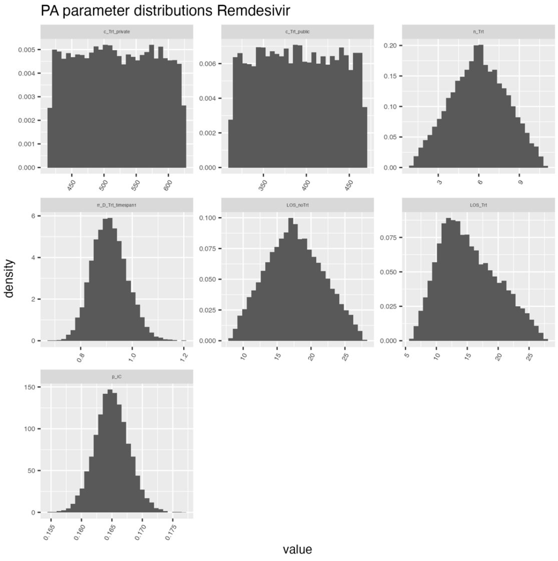

### Tocilizumab

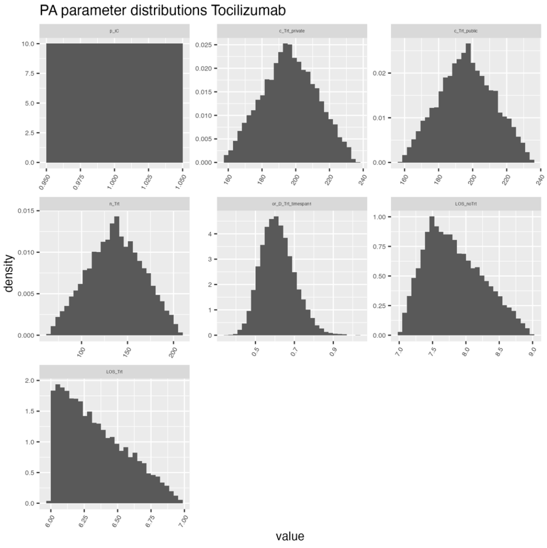
